# Individuals whose phenotype deviates from genetic expectation defined by common variation are enriched for rare damaging variants in genes that cause rare disease

**DOI:** 10.64898/2025.12.30.25343229

**Authors:** Nikolas A. Baya, Frederik H. Lassen, Barney Hill, Samvida S. Venkatesh, Hannah Currant, Cecilia M. Lindgren, Duncan S. Palmer

**Affiliations:** Big Data Institute, Li Ka Shing Centre for Health Information and Discovery, University of Oxford, Oxford, United Kingdom; Centre for Human Genetics, Nuffield Department of Medicine, University of Oxford, Oxford, United Kingdom; Department of Statistics, University of Oxford, Oxford, United Kingdom; Nuffield Department of Population Health, Medical Sciences Division, University of Oxford, Oxford, United Kingdom; Nuffield Department of Women’s and Reproductive Health, Medical Sciences Division, University of Oxford, United Kingdom; Broad Institute of MIT and Harvard, Cambridge, Massachusetts, United States of America; The Pioneer Centre for SMARTbiomed, Big Data Institute, Li Ka Shing Centre for Health Information and Discovery, University of Oxford, Oxford, United Kingdom

## Abstract

Polygenic scores (PGS) predict complex traits and stratify disease risk but often fail to fully capture individual-level variation. “Misaligned” individuals, whose observed phenotypes deviate from their genetically expected values based on polygenic scores (PGS), provide a powerful model for identifying factors beyond common-variant effects, including additional genetic factors. Here, we apply misalignment classification and enrichment testing frameworks to seven continuous and three dichotomous traits, assessing whether misaligned individuals in the UK Biobank are enriched for rare (minor allele frequency (MAF) *<* 0.1%) damaging genetic variation. We identify significant enrichment (false discovery rate (FDR)-adjusted *P <* 0.05) of predicted loss-of-function (pLoF) variants in *COPB2* and *GORAB* among individuals misaligned for lower-than-expected bone mineral density. We refine previously observed grouped-gene enrichment in individuals with misaligned stature to the single-gene level: shorter-than-expected individuals are enriched for pLoF variants in *ACAN* and *IGF1*, and taller-than-expected individuals are enriched for predicted damaging missense in *FBN1*. Using an individual’s misalignment classification as a phenotype, we perform an exome-wide scan across seven traits, resulting in 74 FDR-significant genes. We identify *KANK1* as a gene associated with later age at menopause, potentially protective against primary ovarian insufficiency. For dichotomous disease status traits, we demonstrate evidence for the liability threshold model in the context of counteracting conditionally-orthogonal common and rare variant pathogenic/protective effects. Among individuals diagnosed with type 2 diabetes, carriers of rare pathogenic pLoF variants in *HNF1A* and *HNF4A* had significantly lower polygenic risk than non-carriers (FDR-adjusted one-sided *t*-test *P <* 5 × 10*^−^*^3^). We also show that coronary artery disease controls carrying rare protective pLoF variants in *ANGPTL3* had nominally higher polygenic risk (one-sided *t*-test *P* = 0.03) than non-carriers. This study highlights the power of misalignment-based analyses in complex continuous phenotypes and disease, with the potential to validate known genetic contributors to traits and identify novel genes. This work paves the way for better molecular diagnoses and targeted therapeutic discovery.

## Introduction

Modeling *how* an individual develops disease is key to preventing disease. One approach to modeling genetic disease risk is the liability threshold model^1,2^. This model assumes a continuous “liability” for disease, comprised of genetic and environmental contributions^2^. An individual develops disease if their liability exceeds a certain threshold. The genetic risk portion of this liability can be separated into risk contributed by common variants and rare variants for a heritable trait^3^ (Figure 1).

**Figure 1:**
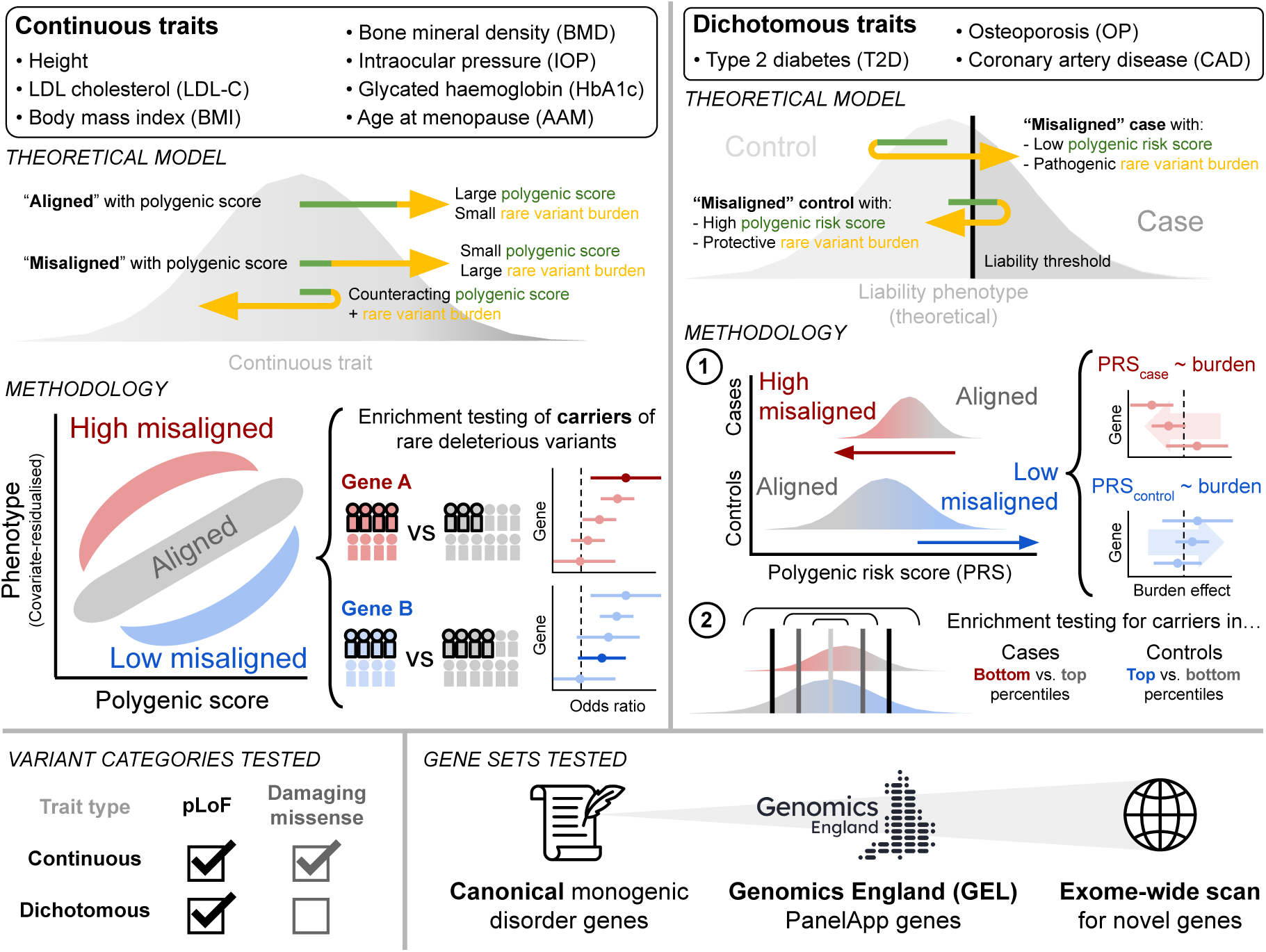
Overview of phenotypic misalignment analysis. We investigated seven continuous and three dichotomous traits to determine if deviation from common-variant genetic prediction is driven by rare damaging variants. **Theoretical Model:** For continuous traits, misaligned individuals are defined by large residuals between observed and expected phenotypes (PGS). For dichotomous traits, under the liability threshold model, misaligned individuals include cases with low PRS) and controls with high PRS, with misalignment driven by pathogenic or protective rare variants. **Methodology:** Enrichment in continuous traits was assessed by comparing rare variant carrier rates in high/low misaligned groups against aligned individuals. For dichotomous traits, we utilized a two-step process: regression of PRS against rare variant burden, and enrichment testing based on PRS percentiles. **Variant categories tested:** We computationally annotated variants and used pLoF or putatively damaging missense variants in our enrichment and burden tests. Only pLoF variants were tested for dichotomous traits. **Gene sets tested:** Tests were applied in stages across canonical monogenic disorder genes, GEL PanelApp gene panel genes, and an exome-wide scan.

One way to quantify the common-variant genetic component of a phenotype is with polygenic scores (PGS)^4,5^. PGS have transformed the study of complex traits by enabling the prediction of phenotypes and disease risk based on the aggregate contribution of common genetic variation^6–9^. However, given the many other contributors to phenotypic variability in the population, assessing how an individual’s observed phenotype deviates from expectation based on their PGS could refine signal for other components (e.g. rare variants, environmental effects) and improve power in association studies. For example, adjusting for PGS has been demonstrated to improve power in rare variant association analyses^10^.

There are other applications for this notion of divergence from the genetically-expected phenotype measured by aggregating common-variant effects. Under a liability threshold model, individuals diagnosed with disease but with low common-variant risk may be expected to have other independent components of risk causing them to cross the liability threshold. This is consistent with recent literature showing that diagnosed individuals with low common-variant risk are more likely to carry rare pathogenic variants^11^ or have a monogenic diagnosis^12^. There are many other recent examples of work involving the interplay of polygenic and monogenic risk, especially demonstrating how common-variant polygenic risk can modulate the penetrance or expressivity of monogenic risk variants. Disorders for which this has been demonstrated include breast cancer^13–15^, Huntington’s disease^16,17^, glaucoma^18,19^, familial hypercholesterolaemia (FH)^13,20^, obesity^20^, developmental disorders^21,22^, chronic kidney disease^23^, and schizophrenia^24^.

Here, we test for enrichment of rare (MAF *<* 0.1%) deleterious variants in UK Biobank (UKB) individuals who deviate from their genetically-expected phenotype based on PGS (Figure 1), using common-variant PGS calculated by genomics PLC^25^ and provided through UKB. We assess seven continuous traits (standing height, low-density lipoprotein cholesterol (LDL-C), body mass index (BMI), glycated haemoglobin (HbA1c), bone mineral density (BMD), intraocular pressure (IOP), and age at menopause (AAM)) and three dichotomous disease status traits (type 2 diabetes (T2D), coronary artery disease (CAD), and osteoporosis (OP)) in our analysis. These traits were selected due to having polygenic and monogenic components of heritability. The selection of dichotomous traits was also motivated by the availability of their corresponding continuous “liability” traits (HbA1c for T2D, LDL-C for CAD, BMD for OP), which enabled us to use many of the same gene sets between traits. For continuous traits, we test for rare putatively deleterious variation in individuals with misaligned phenotypes using an existing framework^26^, and develop a new testing framework to enable the analysis of dichotomous traits.

We build the scope of our investigation in stages. We begin our enrichment testing in a small set of “canonical” genes, curated for each trait, linked to monogenic disorders or strongly associated with the trait. In this context, we have a strong prior for misaligned individuals to show enrichment for deleterious rare variation, given previous work in a smaller subset of UKB^26^.

We then expand our analysis to a larger set of genes considered to be of diagnostic grade for rare disorders related to our selected traits, taken from the Genomics England gene panel^27^. This approach allows us to test many more genes than the canonical genes while maintaining a hypothesis of rare genetic disorder enrichment in misaligned individuals.

Finally, we explore whether this framework of testing for deleterious variant enrichment in misaligned individuals can be re-appropriated for discovery of novel pathogenic or protective genetic effects. To do this, we treat misalignment as the trait of interest and perform gene-level association testing of protective and pathogenic damaging rare variation across the exome.

Altogether this study comprises the largest investigation of phenotype deviation to date, providing further evidence for models of counteracting pathogenic/protective common and rare variant effects.

## Methods

### Exome sequencing data quality control and assigning population labels

We performed sample-, variant- and genotype-level quality control (QC) for the UKB 450k exome sequencing (ES) dataset following the approach of Karczewski *et al.*^28^. Broad continental genetic ancestry labels (Africans, Admixed Americans, East Asians, Europeans, South Asians) were assigned following the approach described by Lassen *et al.*^29^, using a random forest (RF) classifier trained on the 1000 Genomes dataset^30^. We subset to individuals with European genetic ancestry, resulting in a high quality call set of 402,375 samples and 25,229,669 variants.

### Variant consequence annotation

We annotated ES variants using Variant Effect Predictor (VEP) v105 (corresponding to GENCODE v39)^31^ with the Loss-Of-Function Transcript Effect Estimator (LOFTEE) v1.04 GRCh38^32^ and dbNSFP^33^ plugins, annotating variants with Combined Annotation Dependent Depletion (CADD) v1.6^34^, and Rare Exome Variant Ensemble Learner (REVEL) using dbNSFP4.3^35^ and loss-of-function (LoF) confidence using LOFTEE. We provide code and instructions for this step in our VEP 105 LOFTEE repository^36^, which contains a Docker/Singularity container for reproducibility of annotations. Next, we ran SpliceAI v1.3^37^ using the GENCODE v39 gene annotation file to ensure alignment between VEP and SpliceAI transcript annotations. For variant-specific annotations we use “canonical” transcripts. We separated variants by transcript using bcftools +split-vep and filtered to MANE Select^38^ protein-coding transcripts. If the gene lacked a MANE Select transcript we selected the canonical transcript defined by GENCODE v39. Using this collection of missense, pLoF, splice metrics, and annotations of variant consequence on the canonical transcript, we determined four variant categories for gene-based testing.

#### Variant consequence categories

1. High confidence pLoF: High-confidence LoF variants, as defined by LOFTEE^32^ (LOFTEE HC).
2. Damaging missense/protein-altering: At least one of:

a. Variant annotated as missense/start-loss/stop-loss/in-frame indel and (REVEL≥0.773 or CADD≥28.1 or both).
b. Any variant with SpliceAI delta score (DS)≥0.2 where SpliceAI DS is the maximum of the set {DS AG, DS AL, DS DG, DS DL} for each annotated variant (where DS AG, DS AL, DS DG and DS DL are delta score (acceptor gain), delta score (acceptor loss), delta score (donor gain), and delta score (donor loss), respectively).
c. Low-confidence LoF variants, as defined by LOFTEE (LOFTEE LC)
3. Other missense/protein-altering: Missense/start-loss/stop-loss/in-frame indel not categorized in (2).
4. Synonymous: Synonymous variants with SpliceAI DS*<* 0.2 in the gene.

REVEL and CADD score cut-offs are chosen to reflect the supporting level for pathogenicity (PP3) from the American College of Medical Genetics and Genomics and the Association for Molecular Pathology (ACMG/AMP) criteria^39^.

### Phenotype curation

#### Continuous phenotypes

We selected phenotypes among those with “Standard polygenic risk scores (PRS)” scores available in UKB^25^. Standing height (UKB field ID: 50) and LDL-C (UKB field ID: 30780), the two phenotypes analyzed by a previous study of phenotype misalignment^26^, were chosen to benchmark our approach.

The five other continuous traits (body mass index (BMI) [UKB field ID: 21001], estimated bone mineral density (BMD) [UKB field ID: 78], intraocular pressure (IOP) [UKB field IDs: 5262, 5254], glycated haemoglobin (HbA1c) [UKB field ID: 30750], and age at menopause (AAM) [UKB field ID: 3581]) were prioritized for inclusion based on sample size in UKB, heritability, how closely the trait available in UKB matched the trait used to create the PGS, and whether there was prior evidence of monogenic disorders or genes with strong effect contributing to trait variability (Table 2). IOP was calculated as the mean between left (UKB field ID: 5262) and right (UKB field ID: 5254) corneal-compensated measurements. We chose the corneal-compensated measurement method instead of the Goldman-correlated measurement method because it has been reported to be a better indicator for glaucomatous eyes^40^. For AAM, we excluded individuals who responded “Do not know” or “Prefer not to answer”.

**Table 1:** REGENIE burden testing overview. . Burden testing with REGENIE was performed for both continuous and dichotomous trait misalignment testing. In continuous traits, it was only used for the exome-wide scan, while in dichotomous traits it was used for all three gene set categories. We tested both pLoF and putatively damaging missense variants for continuous traits, but only pLoF for dichotomous traits. Misalignment testing in continuous traits treated the misaligned status as a dichotomous variable, with misaligned individuals always encoded as a 1 and aligned as 0. Misalignment testing in dichotomous traits was performed on covariate-residualised PRS, a continuous variable. We use one-sided hypothesis tests throughout: for continuous traits, we always test for higher burden in misaligned individuals; for dichotomous traits, we test for a negative burden association in cases and a positive burden association in controls.

| Trait type | REGENIE used for misaligned phenotype testing in... | Variant consequence burden masks | Dependent variable in regression |  | Hypothesized direction of burden effect |
| --- | --- | --- | --- | --- | --- |
| Continuous | · Exome-wide scan | · pLoF<br>· Damaging missense | Misaligned status [encoding] | High misaligned [1] vs. aligned [0] | Positive (OR>1) |
|  |  |  |  | Low misaligned [1] vs. aligned [0] | Positive (OR>1) |
| Dichotomous (disease status) | · Canonical gene enrichment<br>· GEL gene enrichment<br>· Exome-wide scan | · pLoF | PRS' (covariate-residualised PRS) | PRS' in cases | Negative |
|  |  |  |  | PRS' in controls | Positive |

**Table 2:** Monogenic disorder genes for continuous phenotypes. Genes for each disorder are listed in alphabetical order. See Methods for a description of how genes were selected.

| Trait | Disorder | Effect direction of LoF | Genes |
| --- | --- | --- | --- |
| Standing height | Growth disorder | Negative (pathogenic) | *EZH2, FBN1, NSD1* |
|  |  | Positive (pathogenic) | *ACAN, FGFR3, IGF1, IGF1R, NPR2, SHOX* |
| LDL cholesterol (LDL-C) | Familial hypercholesterolemia | Negative (protective) | *APOB, PCSK9* |
|  |  | Positive (pathogenic) | *LDLR* |
| Body mass index (BMI) | Monogenic obesity | Positive (pathogenic) | *LEP, LEPR, MC4R, PCSK1, POMC* |
| Intraocular pressure (IOP) | Primary open-angle glaucoma | Positive (pathogenic) | *MYOC* |
| Bone mineral density (BMD) | Osteogenesis imperfecta | Negative (pathogenic) | *COL1A1, COL1A2* |
| Glycated hemoglobin (HbA1C) | Maturity-onset diabetes of the young (MODY) | Positive (pathogenic) | *GCK, HNF1A* |
| Age at menopause (AAM) | N/A | Positive (protective) | *CHEK2* |

#### Dichotomous phenotypes

We included three dichotomous disease-status phenotypes: type 2 diabetes (T2D) (International Classification of Diseases, 10th revision (ICD-10) code: E11), coronary artery disease (CAD) (ICD-10 code: I25.1), and osteoporosis (OP) (ICD-10 codes: M80, M81). Case status was defined by ICD-10 diagnoses (UKB field ID: 41270). For each disease, individuals with the disease ICD-10 code(s) were defined as cases, while everyone lacking the disease ICD-10 code, but for whom UKB field ID 41270 was defined, were assigned control status.

#### Polygenic scores

We used PGS determined by genomics PLC^25^ and provided through the UKB (data field IDs in Table S1) for our analyses. We chose to use the “Standard PRS” (UKB category 301) scores instead of the “Enhanced PRS” (UKB category 302) because the Standard PRS category of scores are trained solely using non-UKB data^25^, making them less prone to overfitting to UKB, and as a result, due to their availability for more than three times as many UKB samples compared to the Enhanced PRS.

### Monogenic and rare disorder gene curation

#### Canonical monogenic gene curation

We defined “canonical” monogenic genes as genes which are well-known for causing the most common monogenic disorders associated with the phenotype being studied, based on previous studies.

For standing height and LDL-C we selected the monogenic gene set provided in a study of misaligned individuals in a UKB subset^26^, for both consistency and to check for recapitulation of associations (Table 2). For BMI, we selected the first five monogenic obesity genes discovered^41^: *MC4R*, *PCSK1*, *LEP*, *POMC*, *LEPR*. For bone mineral density we chose the two genes (*COL1A1*, *COL1A2*) where LoF variation is the most common cause of monogenic OP^42^. For intraocular pressure we chose *MYOC*, in which mutations are the most common single-gene cause of primary open-angle glaucoma via increased intraocular pressure^43^. For glycated hemoglobin we chose the genes commonly responsible for maturity-onset diabetes of the young (MODY), the most common monogenic form of diabetes^44^. Within the subtypes of MODY, *GCK* -MODY (MODY2) and *HNF1A*-MODY (MODY3), caused by genetic defects in *GCK* and *HNF1A*, respectively, are the two most common subtypes, accounting for around 80% of MODY cases in the UK^45,46^. Patients in both subtypes can present with hyperglycemia^44,47^, which can be indicated by higher than normal HbA1c levels^48^. For AAM there are no common monogenic disorders. As a proxy, we selected *CHEK2*, the gene with highest strength of association in the burden test for pLoF variants for AAM in Genebass^28^ (pLoF burden *P* =3.75 × 10^−32^). Early menopause can be caused by primary ovarian insufficiency (POI), a partly monogenic disorder^49^. However, there are more than 100 putative monogenic POI genes and in UKB most POI cases are not caused by monogenic mechanisms^49^. For T2D we selected the four MODY genes responsible for the most common subtypes of MODY in the UK: *HNF1A*, *GCK*, *HNF4A*, *HNF1B*^45^. For CAD we selected the same familial hypercholesterolaemia (FH) genes (*PCSK9*, *APOB*, *LDLR*) used for LDL-C, because these monogenic risk genes have been shown to interact with CAD polygenic risk to modulate overall disease risk^13^. Finally, for OP we selected the same osteogenesis imperfecta (i.e. monogenic OP) genes used for BMD (*COL1A1*, *COL1A2*).

#### Rare disorder gene curation

To broaden the scan for enrichment to more genes and objectively define gene sets causing rare disorders, we used the evidence-based, expertly-curated and frequently reviewed Genomics England (GEL) PanelApp resource^27,50^.

When selecting rare disorder gene sets within the GEL PanelApp, we selected disorders that cause extreme, early-onset or monogenic forms of the phenotypes in our analysis. We only included genes rated as “Green” (diagnostic-grade, conservatively defined) in the GEL PanelApp and excluded genes where the mode of pathogenicity was “Other” or started with “Loss-of-function variants (as defined in pop up message) DO NOT cause this phenotype”. This resulted in our final list of diagnostic-grade genes for each selected GEL rare disorder, such that LoF variants in each gene were expected to cause the disorder.

### Misaligned phenotype classification

#### Continuous traits

To determine individuals whose observed continuous phenotypes are aligned or misaligned with their genetically predicted phenotype, we followed Hawkes *et al.*^26^. Briefly, covariates are residualized from the observed phenotype. Mahalanobis distance is calculated for each individual in the bivariate distribution formed by the residualized observed phenotype and the genetically predicted phenotype, and converted to *P* -values^51^. Residualized observed phenotypes are then regressed against genetically predicted phenotypes and residuals are calculated for each individual. We then classify an individual as misaligned or aligned if they satisfied the following conditionsHawkes *et al.*^26^:

### Misaligned individuals

- Mahalanobis *P* -value *<* 0.001, and
- Absolute residual between residualized observed and expected phenotypes *>* 2

### Aligned individuals

- Absolute residual between residualized observed and expected phenotypes *<* 1

In the first residualization step, the covariates were *sex*, *age*, *age*^2^, *sex* × *age*, *sex* × *age*^2^, the first 21 principal components (PCs) (calculated on UKB genotype array data^52^ and provided by UKB), and UKB assessment center. Unlike Hawkes *et al.*^26^, we did not account for statins when adjusting LDL-C for covariates. This simplifying omission is conservative, in that it is more likely to result in false negatives than false positives, as we expect cholesterol- lowering medication to attenuate the misalignment effects of deleterious variants associated with hypercholesterolemia.

#### Case-control dichotomous traits

To contrast individuals with aligned versus misaligned disease status we set percentile thresholds on covariate-residualized PRS distributions (Figure S7). Percentile thresholds were chosen to compare the top versus bottom halves (threshold: 50%), quartiles (thresholds: 25%, 75%), and deciles (thresholds: 10%, 90%). This followed the approach of Lu *et al.*^11^, who compared the top and bottom halves of the PRS distribution using point estimates for pathogenic variant prevalence. We included more extreme thresholds (quartiles, deciles) to assess whether enrichment increased in the extremes of the distribution.

#### Filtering to unrelated individuals

We excluded individuals with a high degree of relatedness (third degree or closer) when defining our aligned and misaligned individuals. Using relatedness estimates (*G*) from a genetic relatedness matrix (GRM) created by Scalable and Accurate Implementation of GEneralized mixed model (SAIGE)^53^ from genotype array data, we filtered to a maximal unrelated set of individuals. We created a greedy algorithm which iterated through a list of individuals (sorted by the number of relatives, with fewest relatives first) with at least one relative (*G >* 2^−3.5^; corresponding to relatives third degree or closer), adding their relatives to the list of individuals for removal. Individuals who had already been added to the list for removal were skipped if encountered later in the iteration. We performed this relatedness filter on each phenotype separately, pre-filtering to individuals with non-missing phenotype and covariate data before applying the relatedness filter, in order to maximize sample size for each phenotype.

### Enrichment testing of carrier status and misaligned status

#### Exact tests on contingency tables

To test whether individuals misaligned with genetic expectation are enriched for a binary variable (e.g. carrier status – whether or not an individual carries one or more damaging variants in a gene), we used Fisher’s exact tests^54^, applied to 2 × 2 contingency tables.

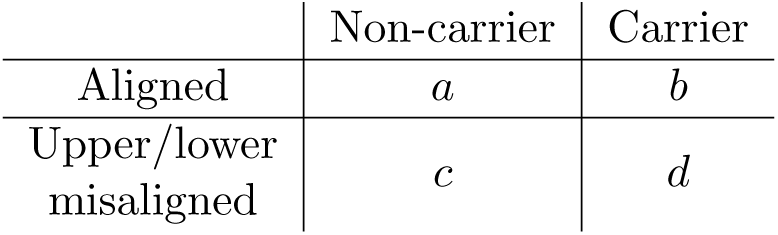

#### Calculating empirical *P* -values with simulated polygenic scores

Spurious misaligned enrichment is possible in our continuous trait misalignment testing framework, whether due to a phenotype effect that is stronger than the misaligned effect, a PGS which explains limited phenotype variability, or a combination of both (Figure S2). To avoid these possibilities, we calculate an empirical *P* -value.

To establish empirical null distributions for enrichment test statistics, we tested the misalignment enrichment method using simulated PGS. The simulated PGS were generated independently of phenotypes, which means the expected phenotypic variance explained is zero. The PGS were simulated from a multivariate standard normal distribution, with zero covariance between individuals.

For each real (using non-simulated PGS) enrichment test in which there were carriers of deleterious variants, we estimated the empirical null by generating replicates of enrichment test statistics based on simulated PGS rather than real PGS (Figure S2). This involved running the misalignment classification procedure using simulated PGS instead of the real PGS, and then testing for enrichment of deleterious variants in the misaligned individuals. The empirical *P* -value was calculated by the following equation:

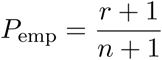

where *n* is the number of test statistic replicates drawn from the null and *r* is the number of those replicate test statistics which are greater than or equal to the observed test statistic^55,56^. We used − log_10_(one-sided Fisher’s exact test *P*) as our test statistic when calculating the empirical *P* -value. We did not exclude replicates where misaligned individuals lacked the variants we were testing; instead, we assigned a Fisher’s exact test *P* = 1.

We simulated 10,000 replicates for all FDR-adjusted significant trait-gene-consequence combinations from enrichment tests of canonical and GEL genes. We simulated 1,000 replicates to test significant genes resulting from the exome-wide scan. We used a reduced number of replicates for the exome-wide scan genes due to the high computational and financial cost when running replicates on the DNAnexus Research Analysis Platform (RAP).

### Hypergeometric test for probability of observing no carriers

When testing misaligned individuals for enrichment of rare (MAF *<* 0.1%) pLoF variants in monogenic disorder genes, there were many instances where, due to the rarity of being misaligned compounded with the rarity of carrying rare pLoF variants, there were no misaligned individuals carrying rare pLoF variants. To quantify the high likelihood of observing no overlap in misaligned status and pLoF carrier status, we used the hypergeometric distribution to calculate the probability of observing no carriers (*k*=0) in a sample of *n* individuals (mis-aligned individuals) from a larger group of *N* individuals (aligned and misaligned individuals) with *K* carriers.

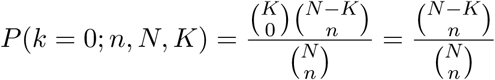

### Burden testing on phenotype misalignment

We used REGENIE version 4.1^57^ to perform gene-level rare variant (MAF *<* 0.1%) burden testing of phenotype misalignment for continuous (for exome-wide scan) and dichotomous traits (for canonical and GEL gene enrichment tests and exome-wide scan), using individuals with European genetic ancestry in the ES 450k tranche of UKB.

In REGENIE step 1, we fit the null model using UKB genotype array data QCed with PLINK^58^ (Filtering flags: `--maf 0.01 --mac 100 --geno 0.1 --mind 0.1`). We included *sequencing tranche*, the tranche in which an individual was exome sequenced, as the only covariate. All other standard covariates (e.g. *age*, *sex*, PCs) had already been regressed out of the observed phenotype (continuous traits) or out of the PRS (dichotomous traits).

After fitting the null model in step 1, we carried out gene-level burden testing in REGENIE step 2 for pLoF (continuous and dichotomous traits) and damaging missense (continuous traits only) variants with MAF *<* 0.1% using the UKB ES 450k tranche data.

To test for an association between rare deleterious variation and misaligned status in continuous traits, we treated misaligned status as a dichotomous phenotype (using the --bt flag in REGENIE). We tested high misaligned vs. aligned and low misaligned vs. aligned separately. When testing misalignment in disease status, we did not apply inverse rank-normal transformation (IRNT) to residualized PRS within REGENIE because the PRS were already normalized.

To compare the statistical power in testing misaligned individuals for gene enrichment to traditional approaches, we ran gene burden tests on the original continuous and dichotomous phenotypes used as input to the misaligned phenotype classification framework. We applied IRNT within REGENIE to all continuous phenotypes using the --apply-rint flag, and used the --bt flag for dichotomous traits.

#### Conditioning on common imputed variants when testing misaligned disease status

When testing rare variant burden effects on common-variant PRS for misaligned disease status, there is a possibility that PRS could correlate with rare variants at a given gene due to linkage disequilibrium (LD). This would lead to spurious burden associations. To conditionally orthogonalize the PRS from rare variant burden, we conditioned on common ( *>* 0.1%) imputed v3 variants located in or within a 1 Mb radius of gene(s) tested (LD in Europeans tends not to extend beyond 1 Mb^30^), with gene start and stop locations in GRCh37 coordinates to match the imputed v3 variants. We used PLINK to QC these variants, following similar filtering thresholds use to select variants which generated the PRS^25^ (PLINK filtering flags: `--maf 0.001 --hwe 1e-10`, variants with INFO score *<* 0.8 excluded). We conditioned on these QCed variants in REGENIE step 2 by passing a gene-specific (or grouped-gene) PLINK pgen file with the flag --condition-file. In our exome-wide scan for misaligned disease status, we performed this conditioning on FDR-significant genes to check robustness of our results.

### Defining significant associations

For each stage of enrichment tests, we applied the Benjamini-Hochberg (BH) procedure to all associations, adjusting for FDR ≤ 5%. Statistically significant associations were defined as having FDR-adjusted *P <* 0.05. As our gene sets (canonical genes, GEL gene panels, exome-wide scan) overlapped, we excluded previously tested genes from consideration when calculating FDR-adjusted *P* -values. After FDR correction, we filtered results on gene-level allele counts, to avoid spurious associations in genes with low allele counts. For continuous trait misaligned status testing, only genes with minor allele count (MAC)≥ 5 in the misaligned group being tested were retained. For dichotomous trait misaligned disease status testing, only genes with alternate allele count (AAC)≥ 10 were retained.

## Results

### Identifying individuals with misaligned continuous phenotypes

Individuals were classified as deviating from their genetically expected phenotype if their observed phenotype, adjusted for covariates, is a significant outlier in the bivariate distribution of phenotype and PGS and if the covariate-residualised observed phenotype does not align with a linear model for observed phenotype and PGS (Methods). We applied this method to seven continuous traits in UKB, with the percentage of misaligned individuals ranging from 0.07% (HbA1c) to 0.12% (AAM)(Figure 1; Table S1; Figure S1).

### Individuals with misaligned continuous phenotypes are enriched for predicted loss-of-function variants in monogenic disorder genes

Following categorization into aligned and misaligned individuals, misaligned individuals can be further split into those whose covariate-residualized observed phenotype is higher-than-expected (“high misaligned”) and lower-than-expected (“low misaligned”), relative to their PGS. In our enrichment tests, we tested for enrichment in one of the misaligned groups (either high or low misaligned) against the aligned group, with the aligned group acting as a control group (Methods; Figure 1).

We are interested in finding genes which significantly contribute to phenotype misalignment. Due to the sparsity of rare deleterious variant carrier status and misaligned status, and their overlap, we wish to control the possibility of spurious associations. Misaligned individuals also tend to have extreme covariate-residualized phenotypes. As the canonical and GEL genes we test are known to associate with the traits tested, we expect carrier status to cluster among individuals with extreme covariate-residualized phenotypes (high or low extremes depending on the effect direction of the gene). Thus, we want to be sure that the misaligned associations we observe are distinct from extreme phenotype associations. Related to this, if a PGS explains limited phenotypic variability, the slope of the phenotype-PGS regression will be close to zero. This would result in phenotype-PGS residuals which directly correspond to the covariate-residualized phenotype. Thus, the misalignment enrichment test would reduce to a test on the phenotype, a similar issue to the overlap between misaligned individuals and individuals with extreme covariate-residualized phenotypes.

To increase confidence that any continuous trait misalignment enrichment is distinct from extreme phenotype association and the PGS explains a sufficient amount of phenotypic variability, we calculated an empirical *P* -value for each putatively significant enrichment result by comparing the observed test statistic (one-sided Fisher’s exact test *P* -value) against an empirical null distribution of test statistics (Methods; Figure S2).

We tested misaligned individuals for enrichment of rare (MAF *<* 0.1%) pLoF variants (see Methods for variant consequence definitions) in genes known to cause monogenic disorders relevant to the misaligned phenotype (Methods).

#### Height and LDL-C

Grouping variants across the monogenic stature disorder genes (Table 2), we observed significant enrichment (FDR-adjusted one-sided Fisher’s exact test *P*_adj_ ≤ 0.05) of pLoF variants in individuals with shorter-than-expected stature (odds ratio (OR) [90% confidence interval (CI)] = 85.7 [49.4-149], *P*_adj_ = 1.13 × 10^−15^) (Figure 2; Table S2). We also tested single genes and observed significant enrichment in individuals with shorter-than-expected stature for *ACAN* (OR [90% CI] = 367 [188-717], *P*_adj_ = 3.02 × 10^−17^), *IGF1* (OR [90% CI] = 1280 [125-13200], *P*_adj_ = 4.08 × 10^−3^), and *SHOX* (OR [90% CI] = 183 [31.4-1070], *P*_adj_ = 0.0148) (Figure 2; Table S2).

**Figure 2:**
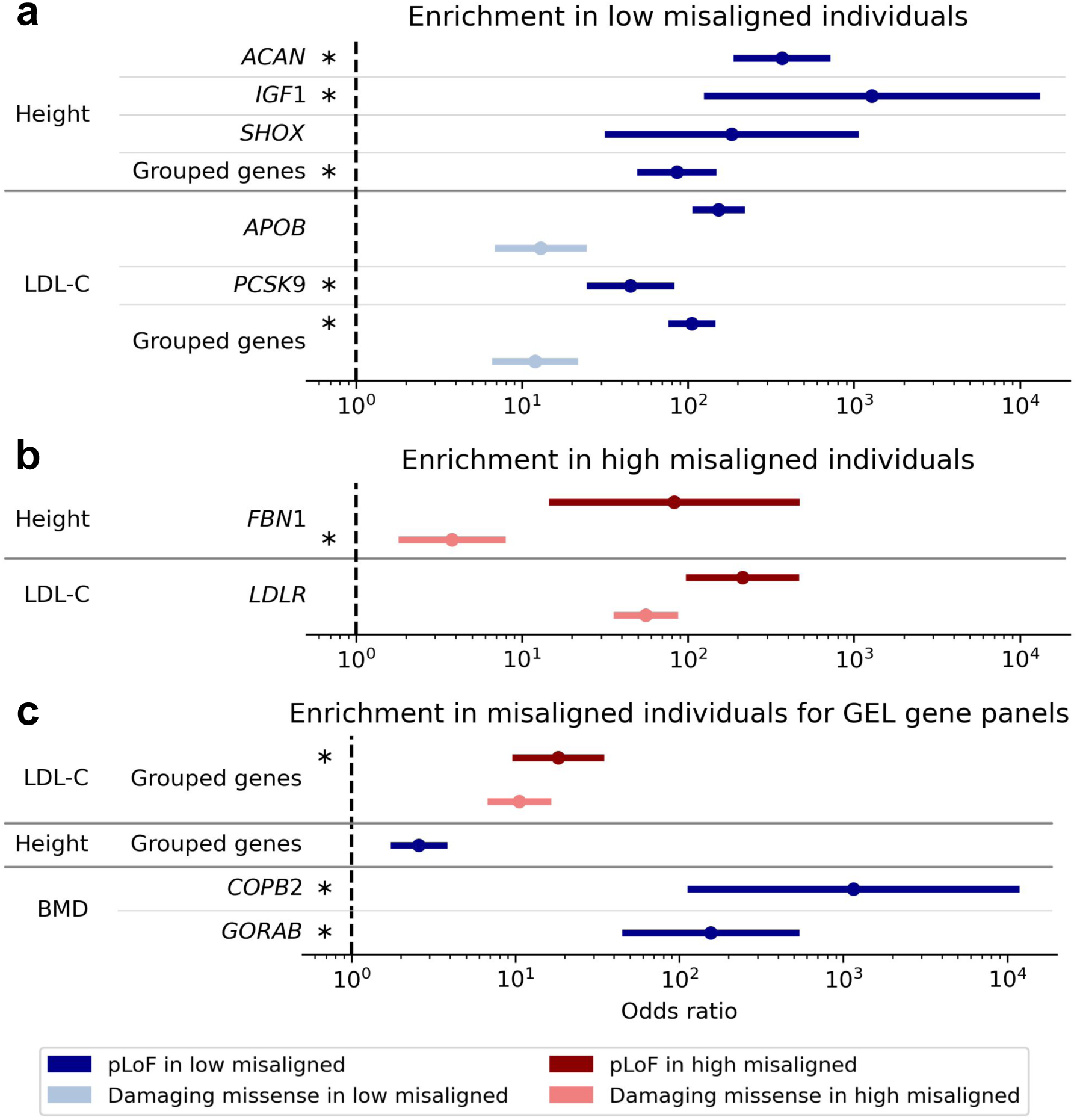
Enrichment of pLoF and damaging missense variants in canonical monogenic disorder and GEL gene panel genes. Enrichment in **a**, low and **b**, high misaligned individuals for monogenic disorder genes and **c**, misaligned individuals for GEL gene panels. “Low misaligned” individuals are individuals with phenotypes lower than genetically expected by PGS. “High misaligned” individuals are individuals with phenotypes higher than genetically expected by PGS. Error bars indicate two-sided 90% CI for OR, such that the lower bound indicates the lower bound for a one-sided 95% CI. Only significant enrichment results (FDR≤ 5%) are shown. Asterisks indicate results where the empirical *P* -value is significant (*P*_emp_ *<* 0.05). Empirical null distributions can be seen in Figure S5.

When testing a set of FH-related genes (Table 2), we found that individuals with high or low LDL-C were enriched for pLoF variants in genes where loss of function is known to cause high (*LDLR* OR [90% CI] = 213 [96.8-468], *P*_adj_ = 3.31 × 10^−10^) or low LDL-C (*APOB* OR [90% CI] = 153 [106-220], *P*_adj_ = 9.45 × 10^−45^; *PCSK9* OR [90% CI] = 45.0 [24.6-82.4], *P*_adj_ = 1.29 × 10^−10^), respectively (Figure 2; Table S2). Individuals with lower than genetically expected LDL-C were also enriched for carrying the combined burden of *PCSK9* and *APOB* pLoF variants (OR [90% CI] = 105 [75.9-146], *P*_adj_ = 5.86 × 10^−51^) (Figure 2; Table S2).

#### Additional continuous phenotypes

For other continuous phenotypes, no misaligned individuals carry pLoF variants in genes causing monogenic disorders (Figure S3; Table 2), likely due to the extremely low frequency of these variants in a population cohort such as UKB. In each trait-gene pair there was a relatively high probability (minimum of hypergeometric test *P* (carriers = 0) = 0.83; Methods) of observing zero pLoF carriers in the set of misaligned individuals, given the number of carriers in the group of misaligned and aligned individuals tested.

Missense variants which are deleterious but not predicted to result in loss of function (“damaging missense” variants; Methods) are by definition expected to have less of an effect on gene product than pLoFs. Assuming a causal link between gene product and phenotype, this implies that damaging missense variants are also expected to have attenuated effect sizes on the phenotype compared to pLoF variants. Although effect size is generally attenuated for damaging missense variants, they tend to occur in higher frequencies. For example, the aggregated pLoF allele count across genes tested for enrichment was 3,114; for damaging missense alleles the count was 17,984, almost six-fold higher. We hypothesized that testing damaging missense variants instead might boost power in enrichment testing by increasing the number of carriers of putatively damaging variation, albeit at smaller effect size, among misaligned individuals.

### Individuals with misaligned continuous phenotypes are enriched for predicted damaging missense variants in monogenic disorder genes

We tested for enrichment of damaging missense variants in misaligned individuals with the same methods as for pLoF variants, for the same set of continuous traits and corresponding monogenic disorder genes.

We observed a significant enrichment (Fisher’s exact test FDR-adjusted *P*_adj_ ≤ 0.05) of damaging missense variants for four of the nine genes/gene sets where misaligned individuals were enriched for pLoF burden (Figure 2; Table S2). Individuals with higher than genetically expected height were enriched for damaging missense variants in *FBN1* (OR [90% CI]= 3.79 [1.80-7.97], *P*_adj_ = 0.0254). For LDL-C, high misaligned individuals were enriched for damaging missense variants in *LDLR* (OR [90% CI] = 55.7 [35.5-87.4], *P*_adj_ = 3.09 × 10^−21^), while low misaligned individuals were enriched for damaging missense variants in *APOB* (OR [90% CI] = 13.0 [6.87-24.6], *P*_adj_ = 5.64 × 10^−6^) and grouped *APOB* and *PCSK9* variants (OR [90% CI]= 12.0 [6.59-21.8], *P*_adj_ = 2.18 × 10^−8^).

As expected, given that we are testing monogenic genes where damaging missense variants are predicted (Methods) to have a less deleterious effect than pLoF variants, the enrichment of damaging missense variants consistently had lower OR point estimates than pLoF variants (4/4 single- and grouped-gene tests with FDR ≤ 5% significant pLoF and damaging missense enrichment, Figure 2).

### Individuals whose continuous phenotype is misaligned with genetic expectation are enriched for predicted damaging variants in genes related to rare disorders

Having demonstrated that misaligned individuals are enriched for both pLoF and damaging missense variation in monogenic genes for continuous traits, we expanded the set of genes analyzed to include genes which cause rare disorders related to the traits analyzed. To do this, we extracted genes from the expertly-reviewed GEL PanelApp gene sets^27,50^ for rare diseases relevant to the phenotypes included in our analysis (Methods). We restricted analysis to genes which qualified for the highest-confidence category (“Green”), designated to be of “diagnostic-grade”^27,50^.

We tested all continuous traits for enrichment of pLoF and damaging missense variants in GEL rare disease genes in misaligned individuals. We used the same enrichment testing framework as described above for the canonical monogenic genes. We excluded 7.69% (38/494) of single- and grouped-gene masks because they had already been included in the enrichment testing of canonical monogenic disorder genes. We performed the BH procedure^59^ to control FDR at 5% across all pLoF and damaging missense enrichment tests for continuous traits.

There were four single- and grouped-gene masks across three traits (height, LDL-C, BMD) for which misaligned individuals were significantly enriched (Fisher’s exact test FDR-adjusted *P*_adj_ ≤ 0.05) for pLoF or damaging missense variants (Figure 2; Figure S4; Table S2).

Individuals with lower than genetically expected BMD were enriched for pLoF variants in *COPB2* (OR [90% CI]=1150 [112-11900], *P*_adj_=0.0325) and *GORAB* (OR [90% CI]=155 [44.5-539], *P*_adj_ = 3.17 × 10^−3^), genes selected from the osteogenesis imperfecta gene panel in GEL PanelApp (Figure 2; Table S2). Both of these genes also demonstrated significant empirical *P* -values (*P*_emp_ *<* 0.05) (Figure S5).

Individuals with higher than genetically expected LDL-C and individuals with lower-than- expected height were also enriched for deleterious variant burden in grouped GEL PanelApp genes. However, these significant enrichments were driven largely by the canonical monogenic genes previously tested, with attenuated odds ratios compared to the results of the grouped canonical monogenic genes (Figure 2; Table S2).

### Individuals whose disease status is misaligned with genetically expected risk are enriched for predicted loss-of-function variants in monogenic genes

We examined misaligned disease status relative to PRS in three common complex diseases: T2D, CAD, and OP. These diseases have been included in previous UKB studies involving PRS and penetrance of pathogenic variants^11,13^. We tested the hypotheses that (1) disease cases with low PRS are enriched for pathogenic rare pLoF variants and (2) disease controls with high PRS are enriched for protective rare pLoF variants.

For each hypothesis, we took a two-step approach. First, within a given disease status group (cases or controls), we used linear regression to test the association of PRS with (pathogenic or protective) pLoF carrier status. Carrier status is a binary variable, defined as whether or not an individual carries at least one pLoF in the gene(s) tested. Second, similar to earlier work in a smaller subset of UKB^11^, we defined low and high PRS percentile groups and tested enrichment of pLoFs between the groups.

To validate our approach, we first applied it to binarized versions of height and LDL-C, continuous traits for which we have already observed an enrichment of rare deleterious variants in individuals whose phenotypes deviate from genetic expectation. Among simulated dichotomized traits with case prevalences of 5% and 10% (similar to the common diseases we aim to study), we replicated all but one (8/9) FDR-significant single- and grouped-gene pLoF associations for misaligned height and LDL-C at nominal significance (one-sided *t*-test *P <* 0.05) (Figure S6; Table S3). This demonstrated proof-of-concept that our pLoF enrichment testing approach for dichotomous case-control traits could recapitulate similar pLoF enrichment in misaligned individuals observed for continuous traits.

#### Disease cases with low polygenic risk are enriched for rare pathogenic predicted loss-of-function variants in monogenic disorder genes

Among T2D, CAD, and OP cases we tested for association of rare (MAF *<* 0.1%) pathogenic pLoF burden with lower PRS. We also tested for association of pLoF burden in low-PRS cases versus high-PRS cases. As with the enrichment testing of misaligned continuous traits, we used a curated set of canonical monogenic disorder genes (Table 3; Methods). Our burden tests were conditioned on common imputed variants in or within 1 Mb of each gene being tested (Methods). We performed the BH^59^ procedure to control FDR at 5% separately for the regression association tests and Fisher’s exact tests.

**Table 3:**
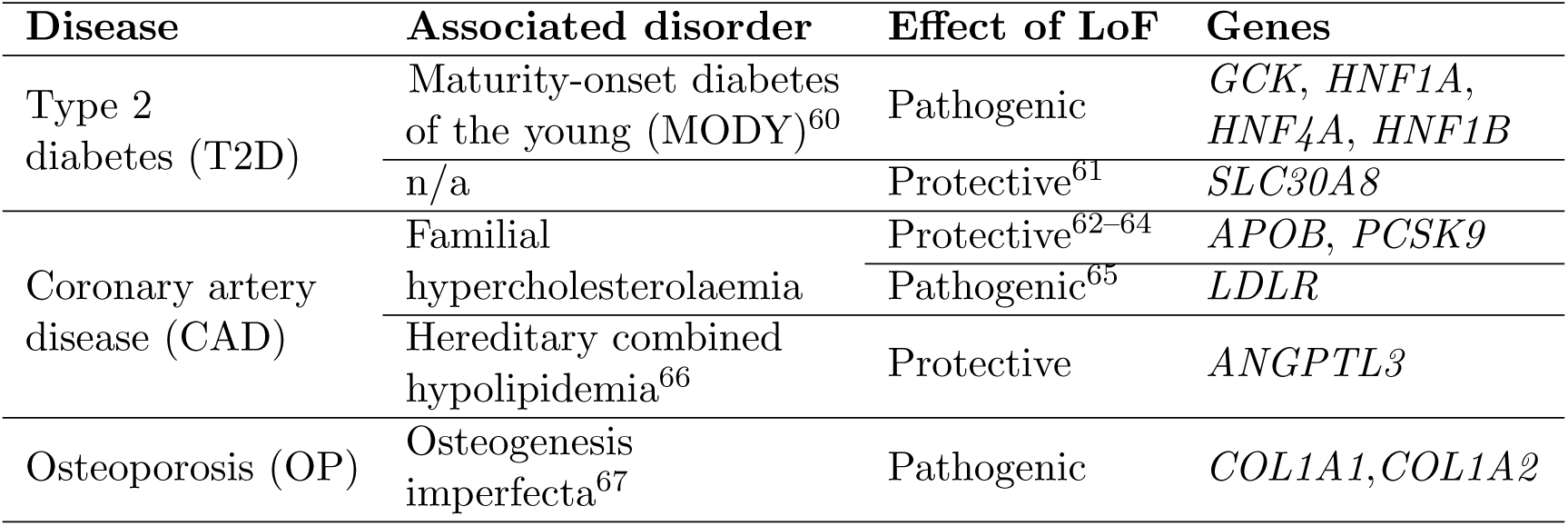
Canonical monogenic genes tested for pLoF enrichment in disease traits.

In T2D cases, pLoF burden was significantly associated (FDR-adjusted one-sided burden *P*_adj_≤ 0.05) with lower PRS for burden in *HNF1A* (*β* [90% CI] = -0.788 [-1.09, -0.484], *P* = 1.03 × 10^−5^, *P*_adj_ = 1.44 × 10^−4^) and *HNF4A* (*β* [90% CI] = -1.88 [-3.08, -0.685], *P* = 4.83 × 10^−3^, *P*_adj_ = 0.0338), with nominally significant (one-sided *P <* 0.05) burden associations in *GCK* and grouped genes (Figure 3a; Table S4).

**Figure 3:**
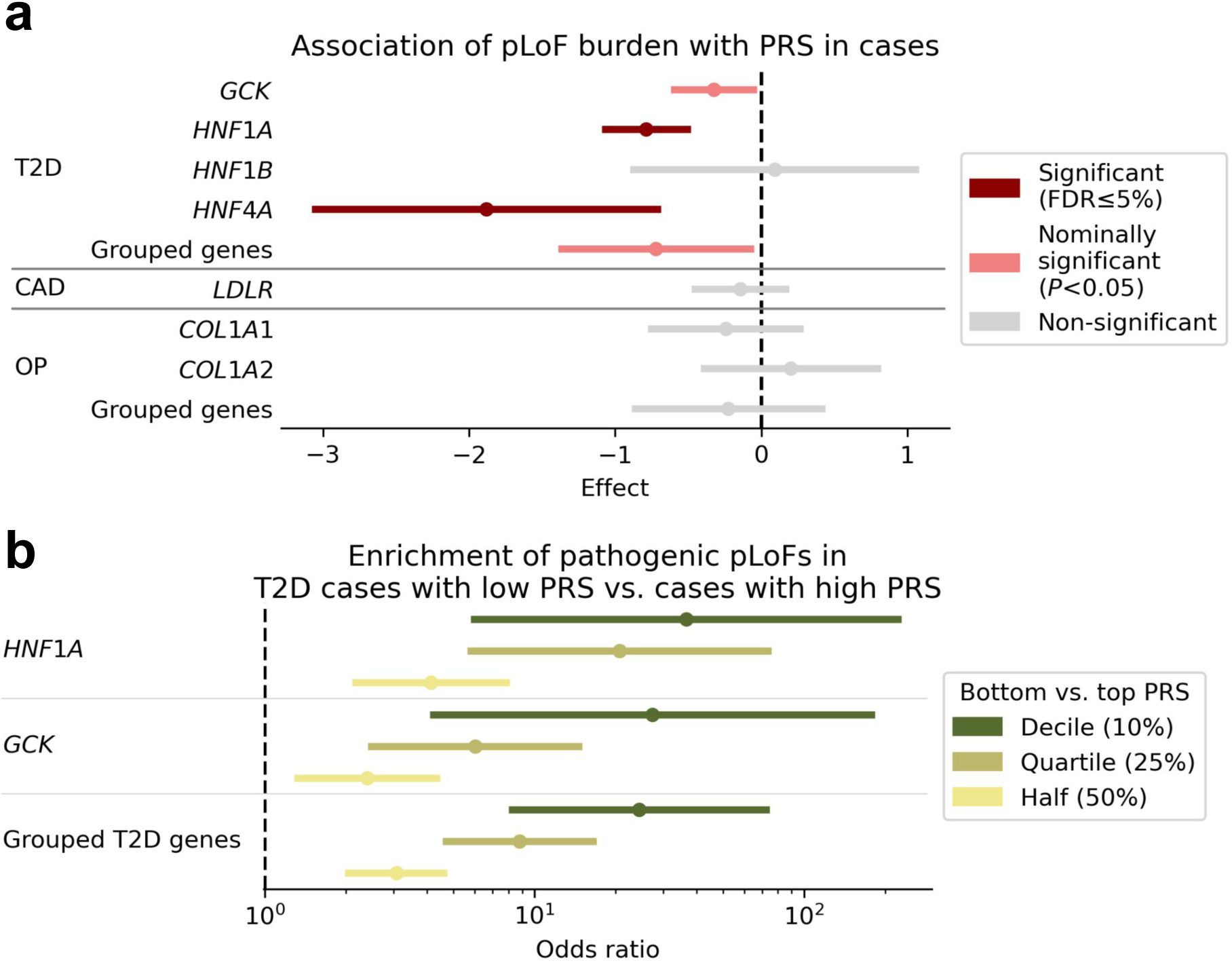
Pathogenic pLoFs are associated with lower PRS in disease cases. **a**, Effect of pLoF variants on PRS among disease cases. Association tests were conditioned on common (MAF *>* 0.1%) imputed variants in or within 1 Mb of each gene tested. **b**, Comparing pathogenic pLoF enrichment in extremes of PRS for disease cases, for genes with significant enrichment results in **(a)**, except for *HNF4A*, which had insufficient carriers to perform enrichment testing in extremes. Error bars indicate two-sided 90% CI for OR, such that the upper bound in **(a)** or lower bound in **(b)** indicates the upper or lower bound, respectively, for a one-sided 95% CI. pLoF, predicted loss-of-function; PRS, polygenic risk score; CAD, coronary artery disease; OP, osteoporosis; T2D, type 2 diabetes.

When testing pLoF enrichment in low vs. high-PRS T2D cases for these single- or grouped- gene masks, we observed a monotonic trend in increasing OR point estimates when comparing ever more extreme PRS quantiles (Figure 3b; Table S5). This suggests that enrichment of pLoFs increases as we compare increasingly misaligned-with-genetic-expectation individuals (low-PRS cases) to increasingly aligned-with-genetic-expectation individuals (high-PRS cases).

#### Disease controls with high polygenic risk are enriched for rare protective predicted loss-of-function variants in monogenic disorder genes

We tested for enrichment of rare pLoF variants in *APOB*, *PCSK9*, and *ANGPTL3* for CAD controls, and *SLC30A8* for T2D controls; these are genes where LoF is expected to be protective for the corresponding disease (Table 3; Figure 4a). Among CAD controls, we observed nominally significant (one-sided burden *P <* 0.05) association of pLoF burden in *ANGPTL3* with increased CAD PRS (*β* [90% CI] = 0.0449 [4.22 × 10^−3^, 0.0856], *P* = 0.0347, *P*_adj_ = 0.108). Comparing relative enrichment of pLoF carriers in disease controls with high versus low PRS, odds ratio point estimates did not demonstrate the same monotonicity observed for pathogenic pLoFs in disease cases (Figure 4b; Table S5).

**Figure 4:**
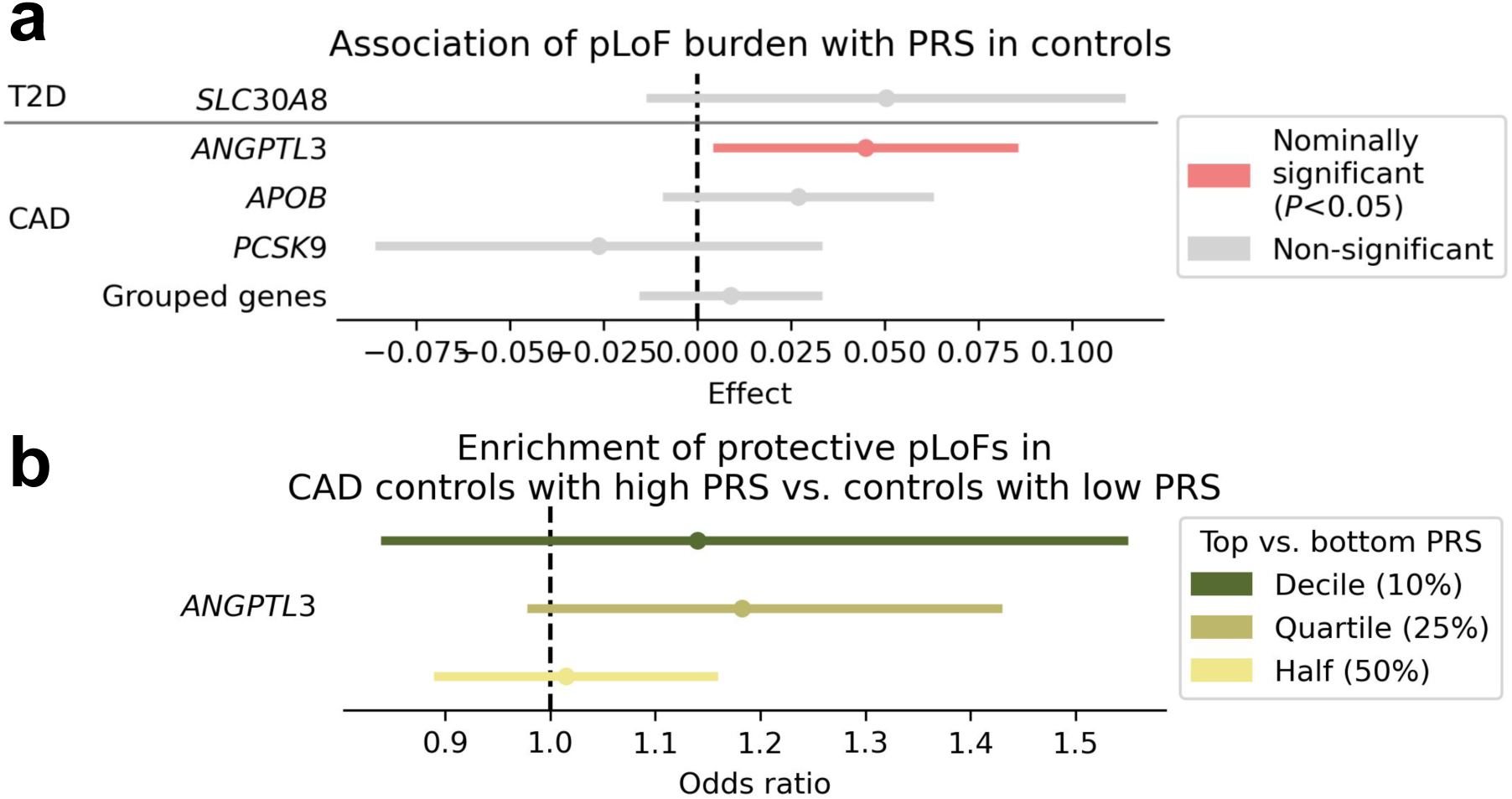
Protective pLoFs are associated with higher PRS in disease controls. **a**, Effect of pLoF variants on PRS among disease controls. Association tests were conditioned on common (MAF *>* 0.1%) imputed variants in or within 1 Mb of each gene tested. **b**, Comparing pathogenic pLoF enrichment in extremes of PRS for disease controls, for genes with significant enrichment results in **(a)**. Error bars indicate two-sided 90% CI for OR, such that the lower bound indicates the lower bound for a one-sided 95% CI. pLoF, predicted loss-of-function; PRS, polygenic risk score; CAD, coronary artery disease; T2D, type 2 diabetes.

#### Limited evidence for enrichment of predicted loss-of-function variants in GEL genes for disease cases with misaligned disease status

We extended our analysis in dichotomous trait misalignment from canonical genes to GEL gene panels. There were no GEL genes where pLoF burden associated significantly (FDR- adjusted one-sided burden *P*_adj_ ≤ 0.05) with lower PRS in cases (Table S4). There were three genes with nominal associations: *BMP1* for OP (*β* [90% CI] = -0.618 [-1.18, -0.0573], *P* = 0.0349), *LMNA* for T2D (*β* [90% CI] = -0.567 [-1.12, -0.0117], *P* = 0.0465), and *LIPA* for CAD (*β* [90% CI] = -0.479 [-0.956, −2.70 × 10^−3^], *P* = 0.0490).

### Exome-wide scan for genes associated with misaligned status in continuous traits

Hypothesizing that there may be genes associated with misaligned status that were not included in the canonical or GEL gene sets, we scanned all genes for deleterious missense variation associated with misaligned status. Genes where deleterious missense variation causes individuals to have lower than genetically expected disease risk, or lower-than-expected liability phenotype, would be ideal targets for therapeutic deactivation. Conversely, a gene where missense variation is pathogenic could be therapeutically targeted for increased expression, assuming druggability of the gene.

For misaligned status of each continuous trait, we performed gene burden testing of rare (MAF *<* 0.1%) pLoF and damaging missense variants with REGENIE^57^, testing separately for protective and pathogenic effects on phenotype misalignment (Methods). Protective effects were defined as those which are trait decreasing, except for BMD and AAM where protective effects are trait increasing. Pathogenic effects are defined as trait-increasing, except for BMD and AAM where they are trait-decreasing. For continuous phenotypes, these definitions are oriented by the diseases of interest for which the continuous phenotypes act as liability phenotypes. The exception was height, where protective effects were defined to be trait decreasing and pathogenic effects trait increasing. These directions were arbitrarily chosen, as there exist both disorders of short and tall stature. When testing misaligned status for protective or pathogenic effects, we always test for enrichment in the misaligned group corresponding to the effect on the original trait (lower-than-expected misaligned when effect is negative, higher-than-expected misaligned when effect is positive).

Burden test *P* -values were controlled using the BH procedure^59^ for FDR≤ 5% across all phenotypes and all genes, separately for protective and pathogenic misalignment tests, with previously tested canonical monogenic disorder and GEL genes excluded. Following FDR control, genes were filtered to those with MAC≥ 5 in the misaligned group being tested, to limit spurious associations. An empirical null *P* -value was calculated for each significant gene enrichment result (Figure S9; Methods).

#### Protective effect enrichment tests

In our exome-wide burden tests for “protective” misaligned status (lower-than-expected misalignment, except for BMD and AAM), 37 genes across six traits harboured significant enrichment (FDR-adjusted one-sided burden *P*_adj_ ≤ 0.05) of pLoF or damaging missense variants (Figure 5a; Table S6).

**Figure 5:**
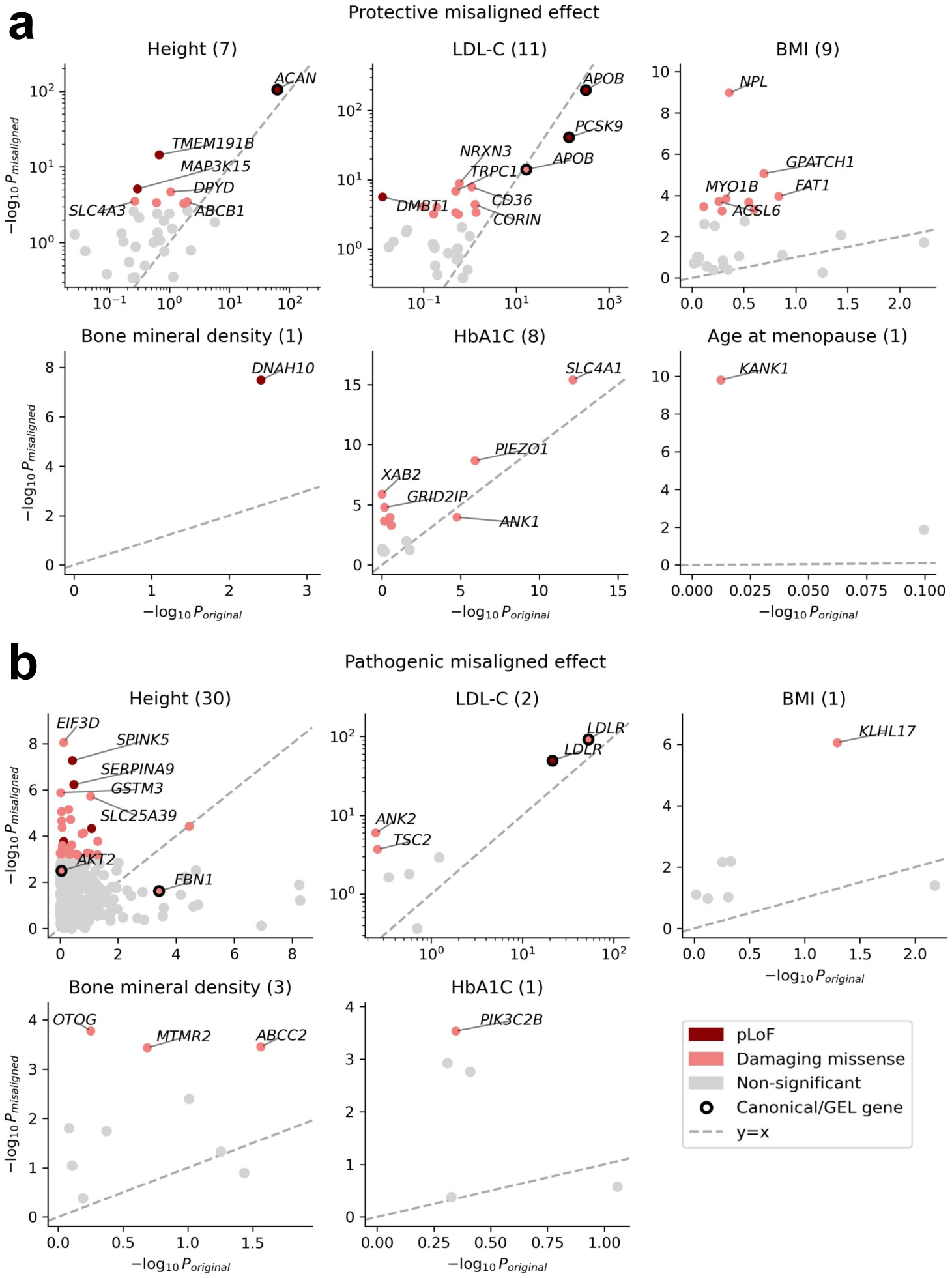
Strength of misaligned burden association, comparing burden test on original phenotype versus burden test of misaligned status. **a**, Protective misaligned effect. **b**, Pathogenic misaligned effect. *P*_misaligned_ is a one-sided *P* -value, with the alternate hypothesis that burden of damaging variants is higher in misaligned individuals (cases) than aligned individuals (controls). *P*_original_ is a two-sided *P* -value for the burden association with the original continuous trait. Only genes with MAC_case_ ≥ 5 are shown. The top 5 FDR-significant genes with highest − log_10_ *P*_misaligned_ are labeled with gene symbol. Canonical and GEL genes are also labeled. The number of FDR-significant genes for each trait is indicated in parentheses in each subplot title. Protective misaligned status corresponds to low misaligned status and pathogenic misaligned status to high misaligned status, except for BMD and AAM, where the effects are flipped (protective=high misaligned, pathogenic=low misaligned).

For lower-than-expected BMI misalignment (9 significant genes), damaging missense associations with plausible biological mechanisms included two genes with links to fat metabolism: *ACSL6* (burden *P*_misaligned_ = 1.99 × 10^−4^, *P*_adj_=0.0182, *P*_emp_ = 0.106; Figure 5a; Table S6), involved in lipid synthesis, such that knockdown in rat skeletal muscle results in decreased lipid accumulation^68^, and *FAT1* (burden *P*_misaligned_ = 1.12 × 10^−4^, *P*_adj_ = 0.0116, *P*_emp_ = 0.0260), involved in fatty acid conversion^69^. The gene with the strongest association, *NPL* (burden *P*_misaligned_ = 1.06 × 10^−9^, *P*_adj_=6.94 × 10^−7^, *P*_emp_ = 9.99 × 10^−4^), has prior experimental evidence supporting a plausible link with decreased BMI. Deficiency of NPL has been shown to result in loss of skeletal muscle in humans, zebrafish, and mice^70^. This loss of muscle mass could lead to lower-than-expected BMI.

For lower-than-expected HbA1c misalignment (8 significant genes), we replicated several known HbA1c associations, including *PIEZO1* ^71^, and two genes (*SLC4A1* and *ANK1*) linked with hereditary spherocytosis^72^, a disorder which can lead to low HbA1c measurements^73^.

Individuals with higher-than-expected AAM were enriched for damaging missense variants in *KANK1* (burden *P*_misaligned_ = 1.55 × 10^−10^, *P*_adj_=1.27 × 10^−7^, *P*_emp_ = 3.00 × 10^−3^; Figure 5a; Table S6). *KANK1* is a uterine fibroid risk gene^74^, much like *CHEK2*, a tumor suppressor gene^75^ where LoF has previously been shown to be associated with increased AAM^76^. Uterine fibroids become less common after menopause^77^, so if damaging variation in a gene was protective for primary ovarian insufficiency, increasing AAM, it might extend the window of time for uterine fibroids to appear, increasing the odds of developing uterine fibroids. When assessing the original trait, association of damaging missense burden in *KANK1* with AAM was null in our study (two-sided burden *P* = 0.964) and Genebass^28^ (missense|low-confidence loss-of-function (LC) burden *P* = 0.879).

#### Pathogenic effect enrichment tests

In our exome-wide burden tests for “pathogenic” misaligned status (higher-than-expected misalignment, except for BMD and AAM), 37 genes across five traits harbored significant enrichment (FDR-adjusted one-sided burden *P*_adj_ ≤ 0.05) of pLoF or damaging missense variants (Figure 5b; Table S6).

For taller-than-expected height, significant enrichment was observed for both pLoF (4 genes) and damaging missense variants (26 genes). Several of the significant genes are known to cause autosomal recessive genetic syndromes which can result in short stature, including *SPINK5* (Netherton syndrome^78^), *PLOD2* (Bruck syndrome^79^), and *GALNS* (Morquio syndrome/mucopolysaccharidosis type IV^80^). These are counter to the direction of effect expected for taller-than-expected height misalignment, however none of the taller-than-expected height outliers were homozygous or compound heterozygous carriers of ClinVar^81^ pathogenic variants from the variant category (pLoF or damaging missense) being tested (Figure S8). This suggests that these carriers are unlikely to have these syndromes, resolving this paradox.

For higher-than expected LDL-C (2 significant genes), one gene with a biologically plausible mechanism was *TSC2* (burden *P*_misaligned_ = 2.01 × 10^−4^, *P*_adj_ = 0.0188, *P*_emp_ = 4.00 × 10^−3^; Figure 5b; Table S6). *TSC2* inhibits the protein kinase mechanistic target of rapamycin complex 1 (mTORC1)^82^; when mTORC1 is left uninhibited, it can promote *de novo* lipogenesis^83^. The other significant gene, *ANK2* (burden *P*_misaligned_ = 1.09 × 10^−6^, *P*_adj_ = 2.96 × 10^−4^), has been strongly implicated in cardiac arrhythmias^84^, but no evidence in the literature suggests that cardiac arrhythmia can cause high cholesterol.

For higher-than-expected HbA1c misalignment, the only significant association was for damaging missense burden in *PIK3C2B* (burden *P*_misaligned_ = 2.93 × 10^−4^, *P*_adj_ = 0.0252, *P*_emp_ = 0.218; Figure 5b; Table S6). There is a plausible pathway implicating this gene in HbA1c, as it encodes a protein involved in the PI3K/AKT/mTOR pathway, a critical regulator of insulin signalling and overall glucose homeostasis^85,86^.

For the remaining traits, there were significant pathogenic damaging missense burden associations for higher-than-expected BMI (*KLHL17*) and lower-than-expected BMD (*OTOG*, *ABCC2*, *MTMR2*). However, these genes did not have strong evidence from existing literature to support their biological plausibility.

## Discussion

In this study, we tested a collection of continuous and dichotomous traits for enrichment of rare deleterious variants in individuals whose observed phenotype deviates from commonvariant genetic expectation. We began with a narrow focus, testing a small curated set of genes canonically implicated in each trait, before expanding to testing larger gene panels from GEL for rare disorders related to the traits. Finally, we broaden our scope to an exome-wide scan, searching for novel gene burden associations with phenotype misalignment.

Using an existing method of misaligned phenotype classification in continuous traits, we replicated many of the pLoF enrichment results previously observed^26^ for misaligned height and LDL-C. We also found that individuals with lower than genetically expected LDL-C and higher than genetically expected stature were enriched for rare damaging missense variants in *APOB* and *FBN1*, respectively. This suggests that phenotype misalignment may be influenced by variants expected to have more moderate effects on gene function. Another novel observation was that in BMD, a trait not investigated by previous studies of misaligned phenotypes, misaligned individuals were enriched for rare pLoF variants in *COPB2* and *GORAB*, genes linked with a monogenic form of OP. These results further validate the use of misalignment classification for prioritizing individuals when screening of rare genetic disorders related to height, LDL-C, and BMD.

Testing for pLoF enrichment in individuals misaligned with disease status, we found that T2D disease cases carrying pathogenic rare pLoFs in certain MODY genes tended to have significantly lower PRS. We also observed that CAD disease controls carrying nominally- significant protective rare pLoFs in *ANGPTL3* tended to have higher PRS. As with the continuous traits, these results support the use of PRS when prioritizing individuals for rare disorder screening. These results also provide strong evidence of a liability threshold model where disease status is determined by a balance of pathogenic and protective effects from common and rare variants (Figure 1).

Moving from validation to discovery, we tested exome-wide for genes associated with protective or pathogenic misaligned continuous phenotype status and identified several genes with plausible biological pathways for their role in causing high or low misaligned phenotypes, including genes associated with lower-than-expected BMI (*ACSL6*, *FAT1*, *NPL*) and higher- than-expected LDL-C (*TSC2*) and AAM (*KANK1*). This demonstrated that misalignment classification of continuous traits could improve power in gene discovery, especially for noisy self-reported phenotypes, such as AAM^87^. We also performed an exome-wide scan for genes associated with misaligned disease status (Supplementary Note 1), but this was less fruitful, yielding a single association in *HLA-DRB5* for CAD-protective pLoF burden. This could be because there are not many genes with effect sizes large enough to counteract common variant effects and cause misaligned disease status.

There were some limitations in this study. First, testing for enrichment of rare damaging variants among misaligned individuals has inherently limited statistical power due to the few or absent variant carriers among misaligned individuals. For example, when testing continuous traits, there were zero variant carriers in misaligned individuals for more than half (32/58) of the canonical monogenic gene enrichment tests and 78.7% (389/494) of GEL PanelApp gene enrichment tests, across all pLoF and damaging missense enrichment tests. This made it impossible to assess relative enrichment of damaging variants in misaligned individuals versus aligned individuals in these instances. In addition to the double jeopardy in statistical power when testing for enrichment of rare variants in a rare group of individuals, there are also expected to be fewer pathogenic variants in UKB because, on average, UKB participants are healthier than the general population^88^. Second, we used variant consequence categories based on computational prediction (see Methods) rather than strictly clinical evidence. Using computational prediction may lead to some miscategorized variants, but enabled us to scale the enrichment tests to many more variants than would be feasible through manual curation of single variants informed by clinical case studies. Third, throughout this paper we use PGS/PRS generated by a single study^25^, provided by UKB. We chose these scores because they were rigorously defined, readily available, and generated independently from our work. Evaluating the performance of our misalignment testing approach across PGS generated by other methods could further generalize our findings and determine what aspects of a PGS make it particularly suited to misalignment testing. Finally, this study was performed solely in individuals classified as having European genetic ancestry (Methods). Identifying misaligned individuals in non-European populations is difficult, as these populations are generally less represented in genome-wide association study (GWAS) with sufficient sample size to generate informative PGS. However, as increasingly diverse large biobanks are created, such as the All of Us (AoU) dataset^89^, misalignment classification will become feasible for more ancestry groups.

There are several areas of opportunity to build off of this work. We could investigate whether non-genetic factors, such as socioeconomic status, diet, or medications, could explain phenotype misalignment. Correcting for these non-genetic factors could also improve power for detecting rare genetic effects on misalignment. Replication of results in an independent cohort, such as the AoU dataset^89^, would further increase confidence in results. Stratifying analysis by sex or another categorical variable and investigating category-specific enrichment, could provide more granularity to the results. However, stratification would increase the multiple testing burden and also decrease the sample size in enrichment tests. Hypothesizing that singletons observed in UKB could be harboring large-effect variants such as *de novo* variants, misaligned individuals could be tested for enrichment of singleton burden. We performed this analysis and found moderate evidence pointing to deleterious singleton burden associating with shorter-than-expected stature (Supplementary Note 2). Singleton burden analysis could be refined by calling *de novo* variants and testing their burden directly, however this was beyond the scope of this project. Unlike the gene-level tests used throughout our work, performing a variant-level exome-wide scan for association could highlight the specific variants driving gene-level associations. However, this higher-resolution approach may only be practical for rare but not ultra rare variants, as statistical power to detect very rare variant associations (e.g. MAC ≤ 10) is attenuated compared to associations in more common variants with the same effect size^90^. Lastly, when performing exome-wide scans for novel protective genetic effects associated with PRS in disease controls, we only tested pLoF burden. This was because we expected pLoFs to have a much stronger effect than other missense variants, with a strong effect presumably necessary to cause disease status misalignment. However, as observed when testing gene burden associated with misaligned status in continuous traits, there may be value in expanding to damaging missense variants.

We demonstrated that for a range of heritable continuous traits and complex diseases, misaligned individuals are enriched for rare damaging variants. This work highlights the value in considering both common and rare genetic components of trait variability, as it may prioritize certain individuals for rare disease diagnosis and could be leveraged to find new pathogenic or protective associations.

## Data Availability

Summary statistics for all analyses not already available in the supplementary materials are available upon reasonable request to the authors.

## Authors

### Correspondence

Correspondence, including requests for further information and resources, should be directed to NAB and DSP.

### Author contributions

Conceptualization: CML, NAB. Funding acquisition: CML. Data curation: NAB, FHL, BH. Methodology: NAB, DSP, SSV, HC. Software: NAB, FHL, BH. Formal analysis: NAB, FHL. Writing – original draft: NAB. Writing – review & editing: NAB, DSP. Visualization: NAB. Project administration: NAB, CML, DSP. Supervision: CML, DSP.

## Resource availability

### Lead contact

Requests for further information and resources should be directed to and will be fulfilled by the lead contact, NAB.

### Data and code availability

Summary statistics for all analyses not already available in the supplementary materials are available upon reasonable request to the authors. All code used in this study will be made available through GitHub upon publication.

## Acknowledgments

This research was conducted using the UK Biobank resource under application number 11867. We thank UK Biobank participants for their contribution.

## Transparent methods

This study used data from the UK Biobank. The North West Multi-centre Research Ethics Committee (MREC) gave approval to UK Biobank as a Research Tissue Bank (RTB) approval. This approval means that researchers do not require separate ethical clearance and can operate under the RTB approval.

## Funding acknowledgments

NAB is supported by the Pembroke College Oxford-Bendich Graduate Scholarship, the Clarendon Fund, and Wellcome Trust Grant Number 224890/Z/21/Z. SSV is supported by the Rhodes Scholarships, Clarendon Fund, and the Medical Sciences Doctoral Training Centre at the University of Oxford. FHL is supported by the Wellcome Trust (award 224894/Z/21/Z) and the Medical Sciences Doctoral Training Centre at the University of Oxford. CML is supported by the Li Ka Shing Foundation, NIHR Oxford Biomedical Research Centre, Oxford, NIH (1P50HD104224-01), Gates Foundation (INV-024200). This research was supported by the Wellcome Trust Core Award Grant Number 203141/Z/16/Z, and Wellcome Trust Residual award (221782/Z/20/Z) with additional support from the NIHR Oxford BRC. The views expressed are those of the authors and not necessarily those of the NHS, the NIHR or the Department of Health. HC is supported by Wellcome (318918/Z/24/Z) and NIH (1P50HD104224-01).

## Supplementary notes

### Supplementary Note 1: Exome-wide scan for genes associated with misaligned disease status

Given the significant associations of pathogenic and protective pLoF burden with PRS among disease cases and controls, respectively, we hypothesized that we could discover novel pathogenic or protective genes by testing exome-wide for PRS association with gene burden.

We performed an exome-wide scan for gene burden associated with protective and pathogenic misaligned disease status, testing burden of rare (MAF *<* 0.1%) pLoF variants. To test for protective burden, we subset to disease controls for T2D, CAD, and OP, and used REGENIE to test for burden associated with increased PRS (Figure 1). When testing pathogenic burden, we subset to disease cases and tested for burden associated with decreased PRS. As a point of comparison, we also performed traditional burden tests on disease status. All tests were one-sided and *P* -values were adjusted for FDR≤ 5% using the BH procedure^59^. FDR correction was performed across results from all phenotypes, but separately for pathogenic and protective tests. There were no FDR-significant associations for pathogenic burden associated with decreased PRS in disease cases (Figure S11; Table S7), thus only the protective burden associations are presented here.

There were two genes where rare (MAF *<* 0.1%) pLoF variant burden was significantly associated (FDR-adjusted one-sided burden *P*_adj_ ≤ 0.05) with increased PRS in controls for CAD (*HLA-DRB5*) and OP (*NBPF3*) (Figure S10, Table S7). After conditioning on common imputed variants within 1 Mb of the gene, the burden association in *HLA-DRB5* remained nominally significant (one-sided burden *P* = 3.89 × 10^−4^).

The gene *HLA-DRB5* (major histocompatibility complex, class II, DR beta 5) codes for the beta chain in a human leukocyte antigen (HLA) class II cell surface complex, involved in generating immune responses^91^. Although CAD is linked with immune response in the context of inflammation from atherogenesis^92^, there is no evidence in the literature of a plausible connection between *HLA-DRB5* LoF and reduced risk of CAD onset.

### Supplementary Note 2: Individuals with genetically unexpected phenotypes are nominally enriched for consequence-stratified singleton burden in intolerant genes

#### Methods for singleton burden association

Singleton variants were defined as variants with an alternate allele count of 1 and alternate allele frequency *<* 0.01%. We used PLINK^58^ to aggregate singleton counts for each chromosome and each variant consequence category. We aggregated singleton burden across all genes predicted to be intolerant to LoF variants. We defined intolerant genes using the gene intolerance threshold (loss-of-function observed/expected upper bound fraction (LOEUF)*<* 0.6) proposed by Karczewski *et al.*^32^, using Genome Aggregation Database (gnomAD) v4.1 constraint metrics downloaded from the gnomAD website. Of the 18,623 canonical transcripts available, 3,900 were defined as intolerant. Subsetting to intolerant genes is expected to increase signal-to-noise ratio when testing variant effects, because variants in tolerant genes are not expected to affect the function of the genes, and by extension, not expected to affect the trait. This filtering is particularly important for exome-wide aggregated burden because there are many more variants being aggregated together than in the previous single- or grouped-gene tests.

We tested for the effect of consequence-specific singleton burden (*s_c_*, for consequence category *c*) using regression, controlling for overall singleton burden by including as a covariate the total singleton burden (*s*_total_) across consequence categories, per Genovese *et al.*^93^. We used logistic regression when testing misaligned status in continuous traits, and linear regression when testing for effect of singleton burden on covariate-residualised PRS (PRS^′^) in cases and controls for dichotomous case-control traits:

|  |  |  |
| --- | --- | --- |
| | Continuous traits | Misaligned status $\sim s_c + s_{\text{total}} + 1$ |
| Dichotomous traits | Cases | PRS'_{\text{cases}} $\sim s_c + s_{\text{total}} + 1$ |
| | Controls | PRS'_{\text{controls}} $\sim s_c + s_{\text{total}} + 1$ |

We calculated one-sided *P* -values for the singleton burden term, given our hypothesis that ultra-rare variants are more likely to cause individuals to be misaligned rather than aligned. We stratified the analysis across four protein-coding variant consequence categories: pLoF, damaging missense, other missense and synonymous (see Methods for variant consequence definitions).

#### Monotonic point estimates of singleton enrichment in height suggests deleterious singleton burden is associated with shorter stature

Among continuous traits, there were no significant (FDR-adjusted one-sided *t*-test *P*_adj_ ≤ 0.05) enrichment results (Figure S12; Table S8) after FDR correction. Approaching the results qualitatively, one notable observation was the monotonic trend across functional categories in enrichment effect sizes for height (Figure S12), a trait that showed strong enrichment when testing deleterious burden in monogenic and rare disorder genes. Individuals with lower- than-expected height were increasingly enriched for more deleterious singleton burden, while individuals with higher-than-expected height were increasingly depleted for more deleterious singleton burden. This suggests that, in general, deleterious singletons are more likely to decrease rather than increase height. This is consistent with the work of Backman *et al.*^94^, who found that, among genes where pLoF or likely deleterious missense singleton burden was significantly associated with standing height, the burden effects in most (8/10) genes decreased height (see Backman *et al.*^94^ Supplementary Data 2). This also aligns with a recent analysis of UKB whole-genome sequencing data^95^, in which four genes where pLoF singletons were significantly associated with height, three genes were associated with decreased height (see Hawkes *et al.*^95^ Supplementary Data 6).

#### Limited evidence for association of singleton burden with misaligned disease status

We performed one-sided *t*-tests for association of singleton burden with PRS for CAD, OP, and T2D. In cases we tested whether higher singleton burden was associated with lower PRS, and in controls we tested whether higher singleton burden was associated with higher PRS. This was based on the same hypotheses used for the canonical and GEL tests: Association of variant burden with lower or higher PRS could indicate effects which are pathogenic or protective, respectively, against common-variant risk scores.

After FDR correction, there were no significant associations (FDR-adjusted one-sided *t*-test *P*_adj_ ≤ 0.05) (Figure S13; Table S9). In CAD cases, effect size point estimates monotonically decreased with consequence deleteriousness, with pLoF and damaging missense (the two most deleterious consequences) singleton burden being nominally associated (one-sided *t*-test *P <* 0.05) with lower PRS. This suggests that deleterious singleton burden in intolerant genes could be pathogenic for CAD. We also observed similar monotonicity in effect size point estimates for T2D cases, suggestive of deleterious burden in intolerant genes being pathogenic for T2D, however, none of the enrichment tests were significant.

## Supplementary figures

**Figure S1:**
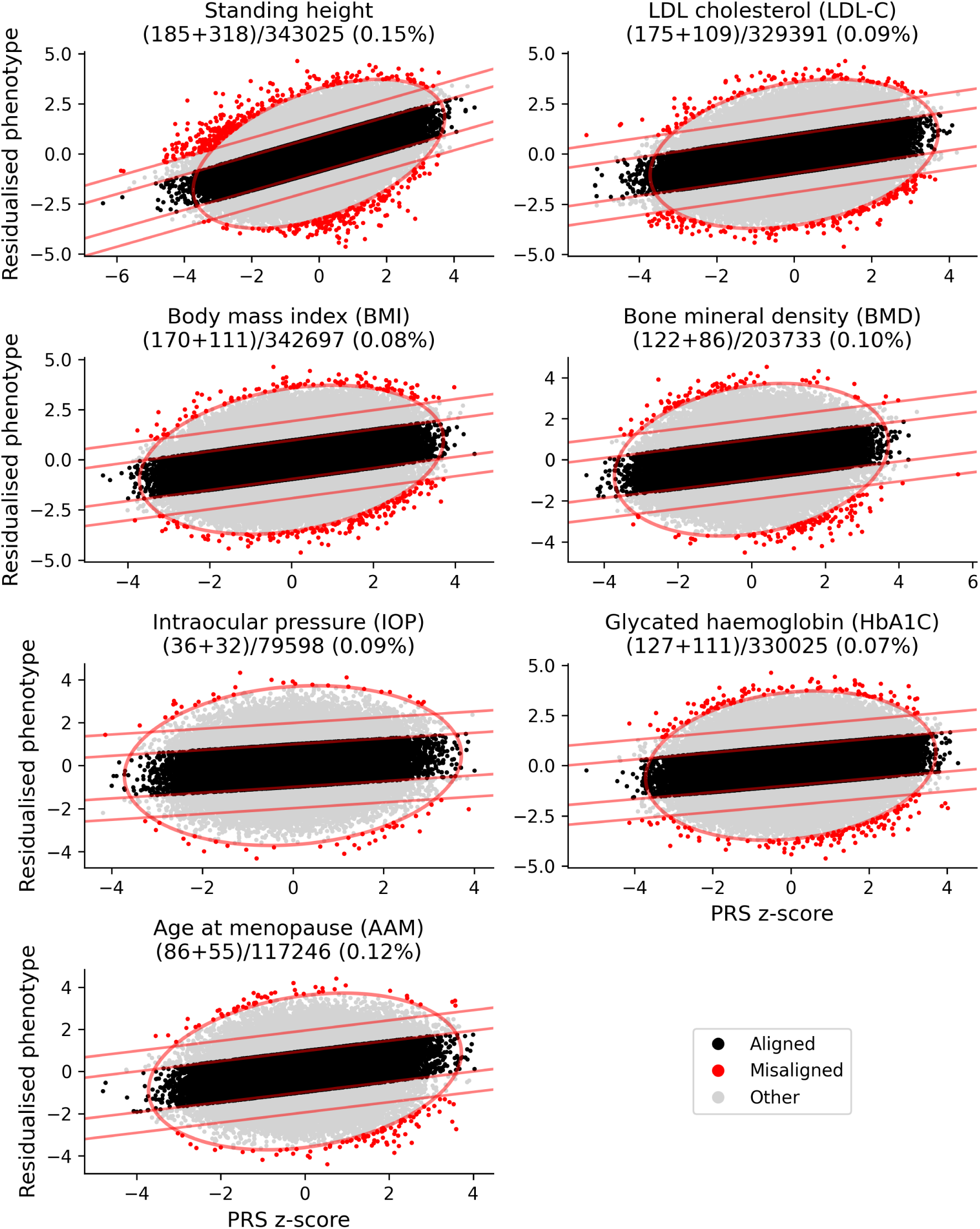
Classification of misaligned individuals for seven continuous traits. In the title of each plot is the name of the phenotype, the sum of low and high misaligned individuals divided by the total number of individuals with defined phenotype, and the total percentage of individuals who are classified as misaligned. The red oval delineates the Mahalanobis distance threshold, while the red lines indicate thresholds on the residuals to define aligned and misaligned individuals.

**Figure S2:**
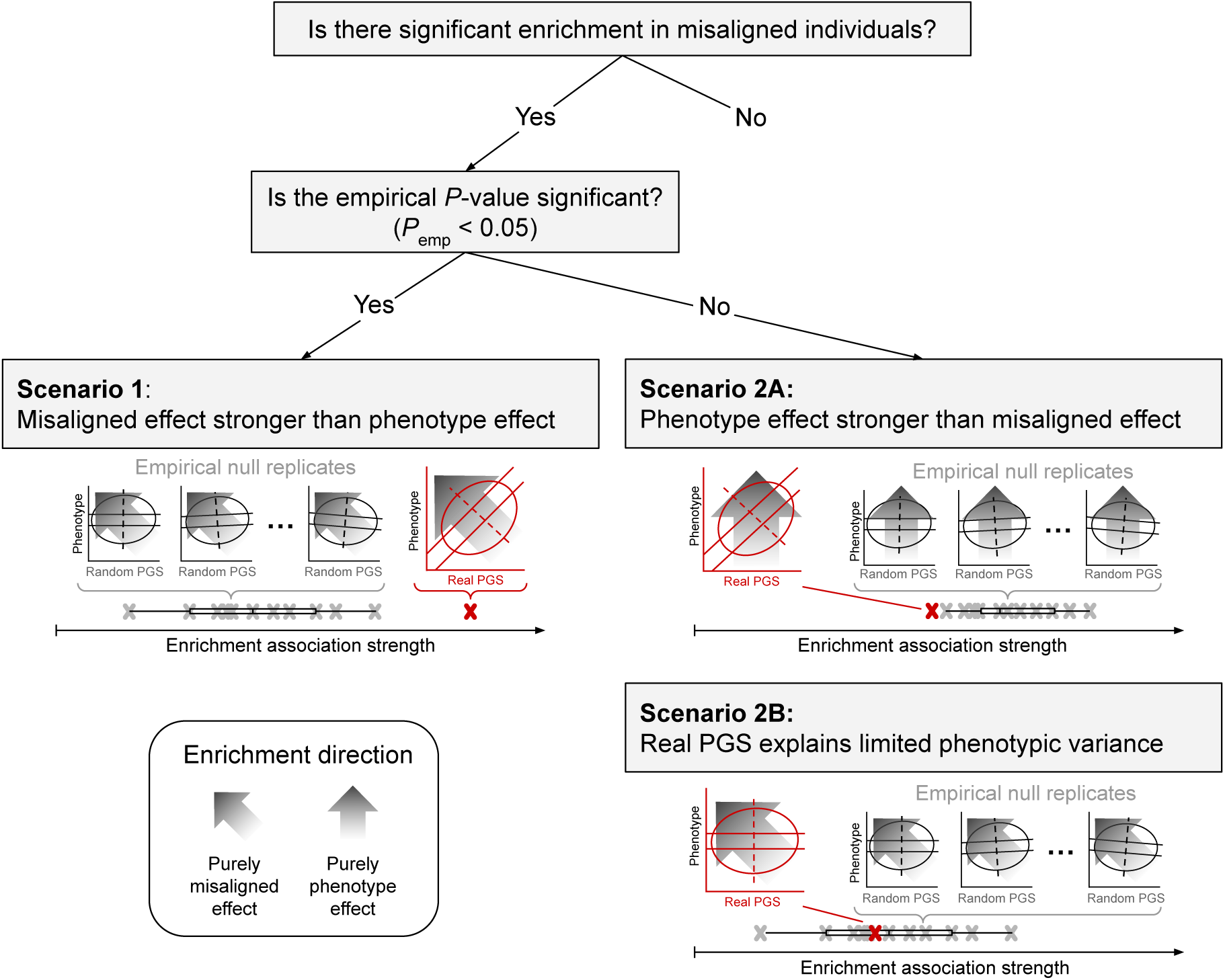
Possible scenarios for the empirical *P* -value. First, note that the empirical *P* -value is only calculated for genes which demonstrate significant enrichment of rare deleterious variants among misaligned individuals. The empirical *P* -value may either be significant or non-significant. To help visualize misaligned enrichment, the gradient of rare deleterious variant enrichment among individuals is shown here as an arrow with gradient shading. The “misaligned axis”, orthogonal to the regression of observed phenotype and PGS, is shown as a dashed line. In this schematic, the alignment of the dashed misaligned axis with the enrichment gradient arrows suggests a strong enrichment. The wider the angle between these two, the weaker the enrichment. **Scenario 1** provides an example of a significant empirical *P* -value, where the observed test statistic (red “×”), based on misaligned enrichment testing using real PGS, outperforms empirical null test statistics (grey ×’s, with box-and-whisker plot for their distribution) generated from misaligned enrichment testing using simulated random PGS. **Scenarios 2A/B** describe examples where spurious enrichment in misaligned individuals can occur, but can be controlled with empirical *P* -values. Note that in reality, the effects described by these scenarios may be more subtle. It is also possible for a mix of both scenarios to occur. **Scenario 2A** describes the possibility that enrichment of individuals is stronger along the observed phenotype axis, rather than along the misaligned axis (orthogonal to the regression of observed phenotype and PGS). Here, empirical null enrichment tests are expected to outperform the real test statistic because, with the random PGS, the enrichment gradient (arrow) is closely matched with the misaligned axis (dashed line). This results in a non-significant empirical null *P* -value. **Scenario 2B** describes the possibility of a real PGS explaining limited phenotypic variance, such that the real test statistic is indistinguishable from the empirical null distribution, resulting in a non-significant empirical *P* -value.

**Figure S3:**
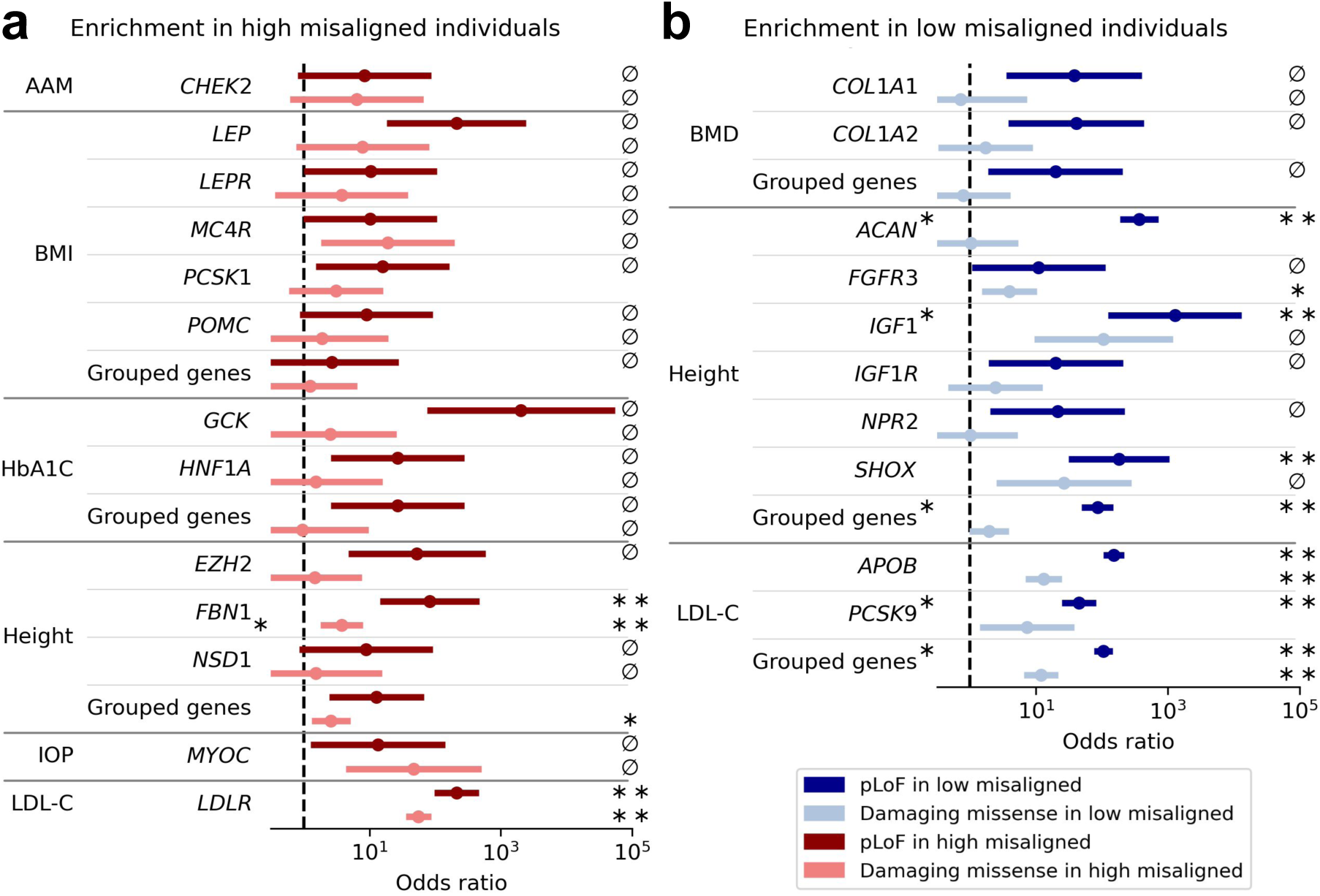
Enrichment of deleterious variants in canonical monogenic genes for continuous traits. **a**, Enrichment testing in high misaligned individuals, in genes where deleterious variants are expected to cause higher-than-expected phenotypes. **b**, Enrichment testing in low misaligned individuals, in genes where deleterious variants are expected to cause lower-than-expected phenotypes. Symbols to the right of each forest plot indicate whether the enrichment test was significant (^∗^*P <* 0.05, ^∗∗^*P*_adj_ *<* 0.05) or whether misaligned individuals lacked carriers (∅) to be able to perform Fisher’s exact test on a fully non-zero contingency table. Asterisks immediately to the right of the gene symbol indicate tests where the observed enrichment outperformed the empirical null (*P*_emp_ *<* 0.05). Odds ratios are calculated for all enrichment tests (with zeros in contingency tables filled with 0.5) and plotted with 90% confidence intervals, such that the lower bound corresponds to the lower bound of a one-sided 95% confidence interval.

**Figure S4:**
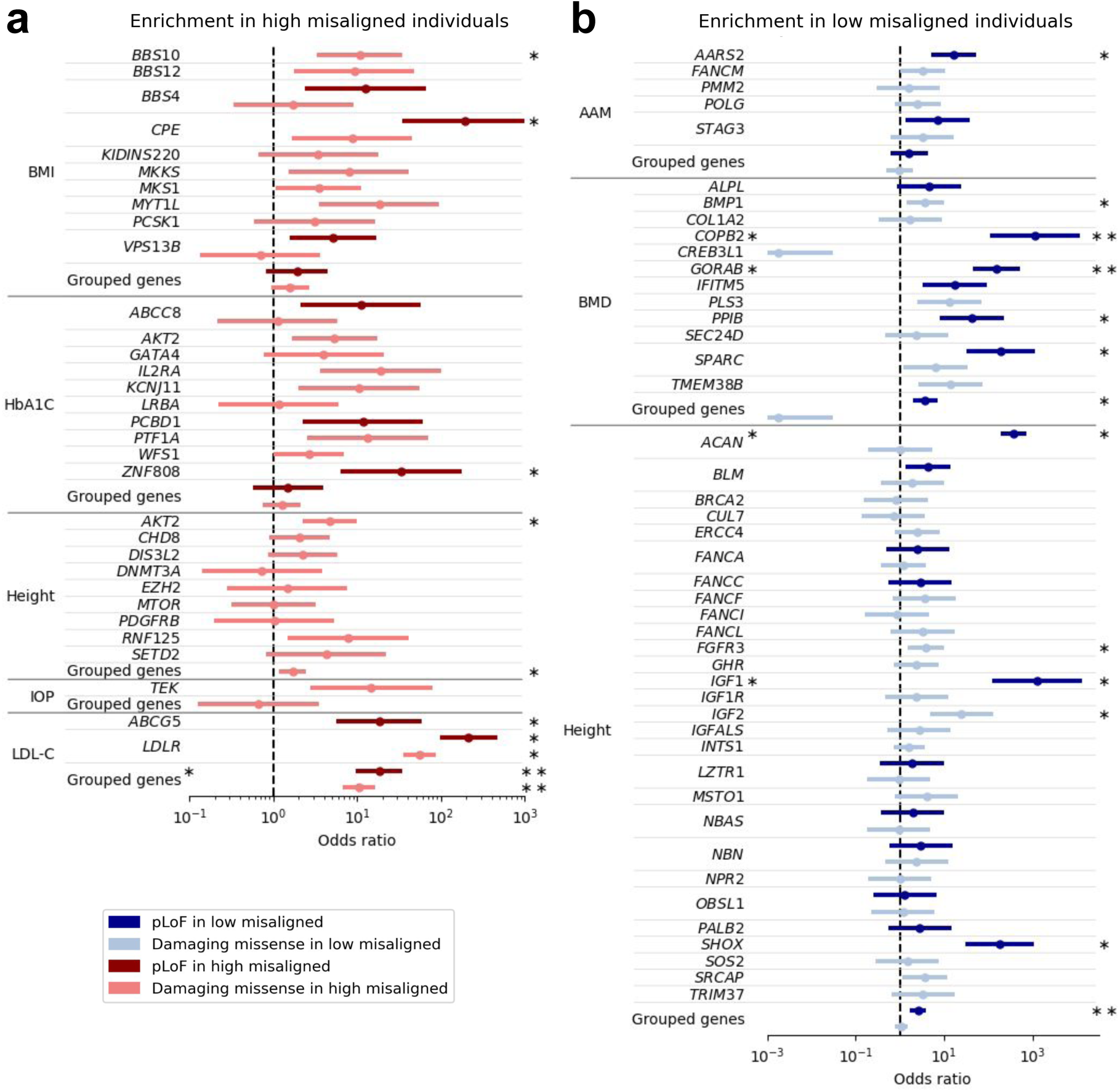
Enrichment of deleterious variants in GEL genes for continuous traits. **a**, Enrichment testing in high misaligned individuals, in genes where deleterious variants are expected to cause higher-than-expected phenotypes. **b**, Enrichment testing in low misaligned individuals, in genes where deleterious variants are expected to cause lower-than-expected phenotypes. Asterisks to the right of each forest plot indicate whether the enrichment test was significant (^∗^*P <* 0.05, ^∗∗^*P*_adj_ *<* 0.05). Asterisks immediately to the right of the gene symbol indicate tests where the observed enrichment outperformed the empirical null (*P*_emp_ *<* 0.05). Odds ratios are calculated for all enrichment tests and plotted with 90% confidence intervals, such that the lower bound corresponds to the lower bound of a one-sided 95% confidence interval. Only enrichment results with fully non-zero contingency tables are shown.

**Figure S5:**
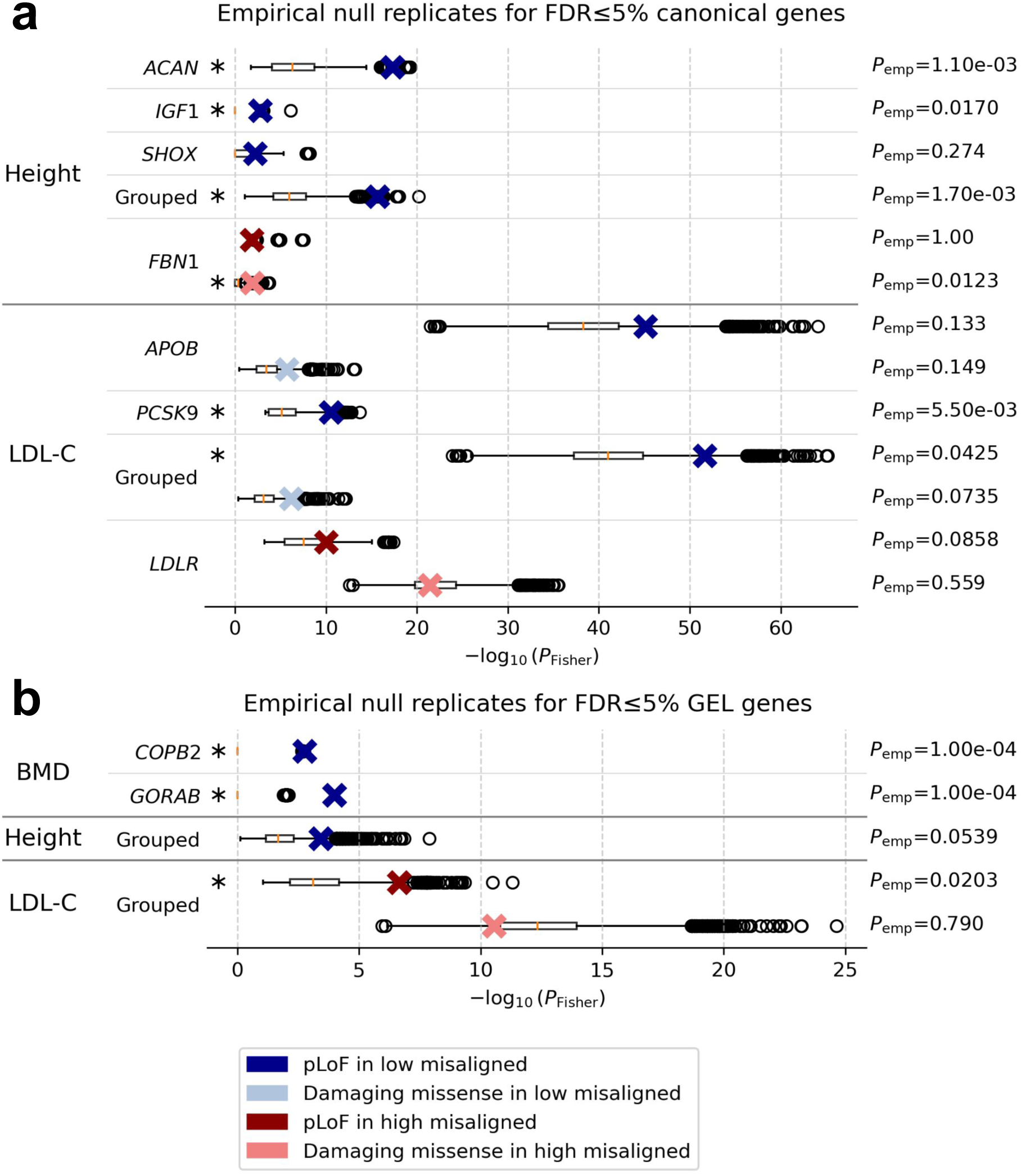
Empirical null replicates for canonical and GEL genes. **a**, Canonical genes. **b**, GEL genes. The empirical *P* -value (*P*_emp_, labeled on the right of each plot) of a gene enrichment result indicates how the observed test statistic (one-sided Fisher’s exact test *P* -value) compares against the empirical null distribution of test statistics (Methods). Asterisks immediately to the right of the gene symbol indicate tests where the observed enrichment outperformed the empirical null (*P*_emp_ *<* 0.05).

**Figure S6:**
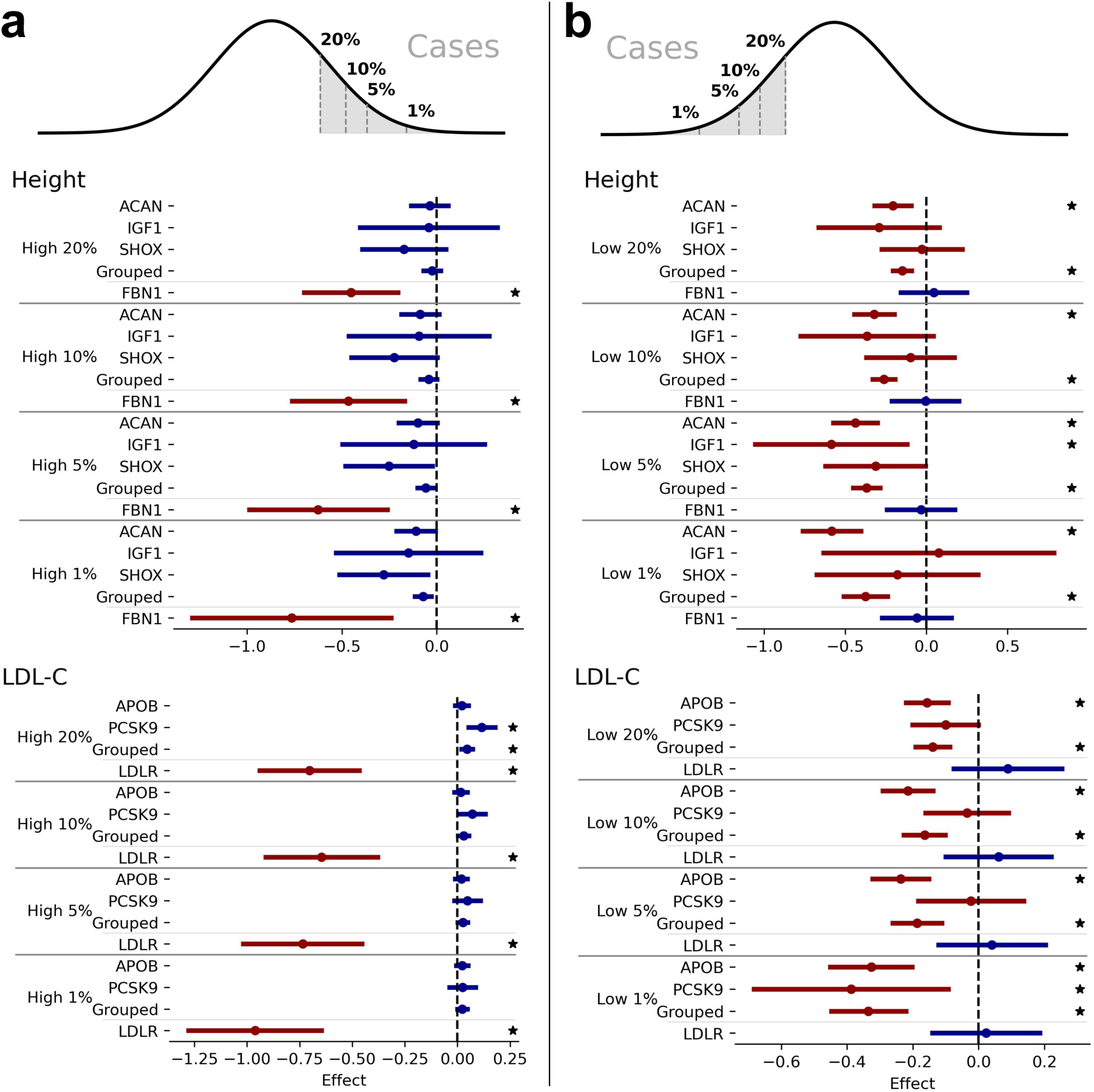
Enrichment testing in FDR-significant canonical genes for simulated dichotomous traits. **a**, Dichotomous traits simulated with the upper tail as cases and everyone else as controls. **b**, Dichotomous traits simulated with the lower tail as cases and everyone else as controls. Only genes and grouped genes which are significant (one-sided Fisher’s exact test *P*_adj_ *<* 0.05) from misaligned enrichment testing of canonical genes in continuous traits are shown. Stars to the right indicate nominal significance (one-sided *t*-test *P <* 0.05) resulting from regression of PRS on carrier status.

**Figure S7:**
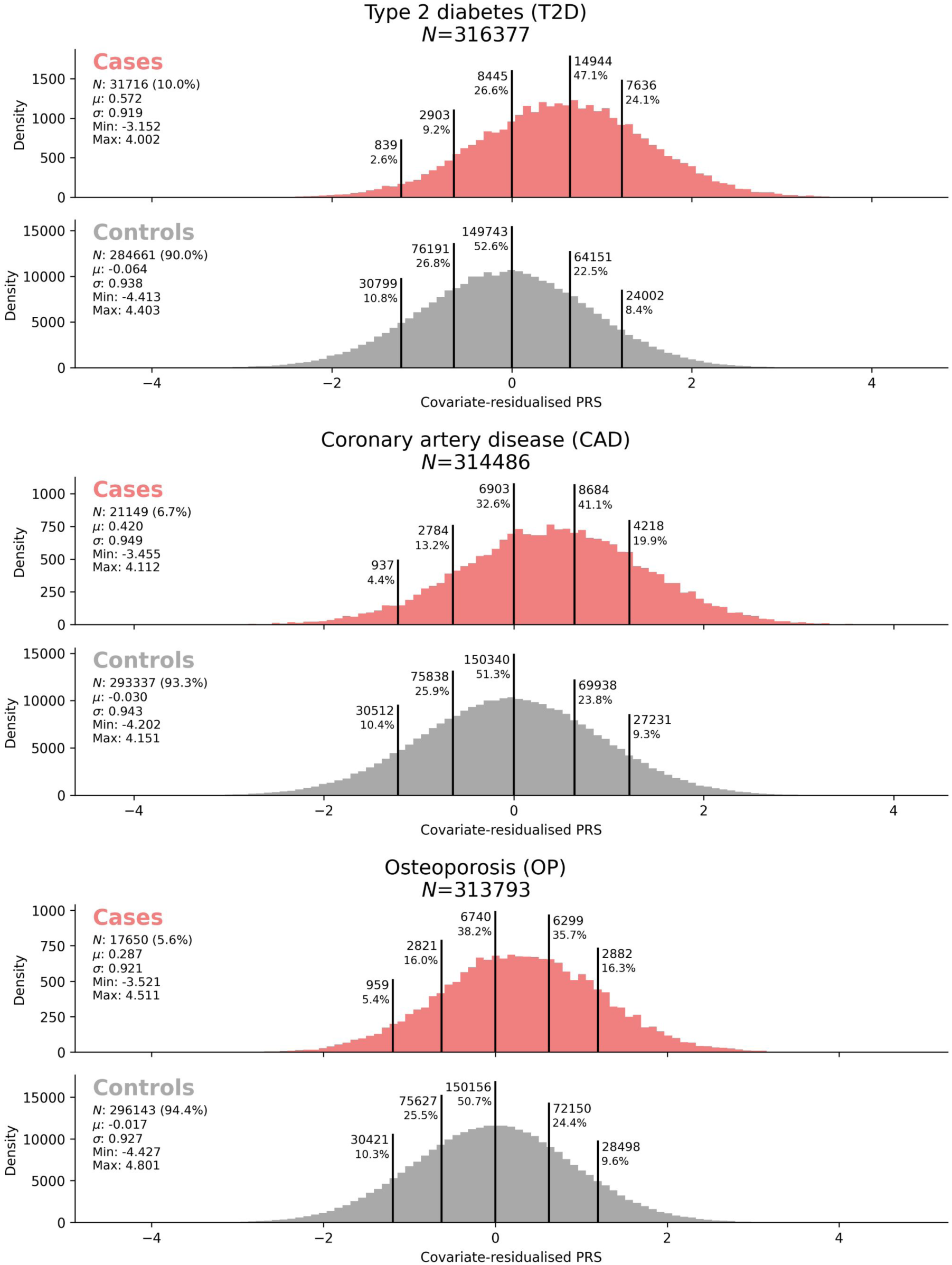
Distributions of covariate-residualised PRS stratified by case status. Vertical lines indicate 10th, 25th, 50th, 75th and 90th PRS percentiles, calculated on the combined distribution of PRS in both cases and controls. Numbers at the top of vertical lines, to the left or right, indicate the number and percentage of samples in the case/control group that lie to the left or right of the vertical line, respectively.

**Figure S8:**
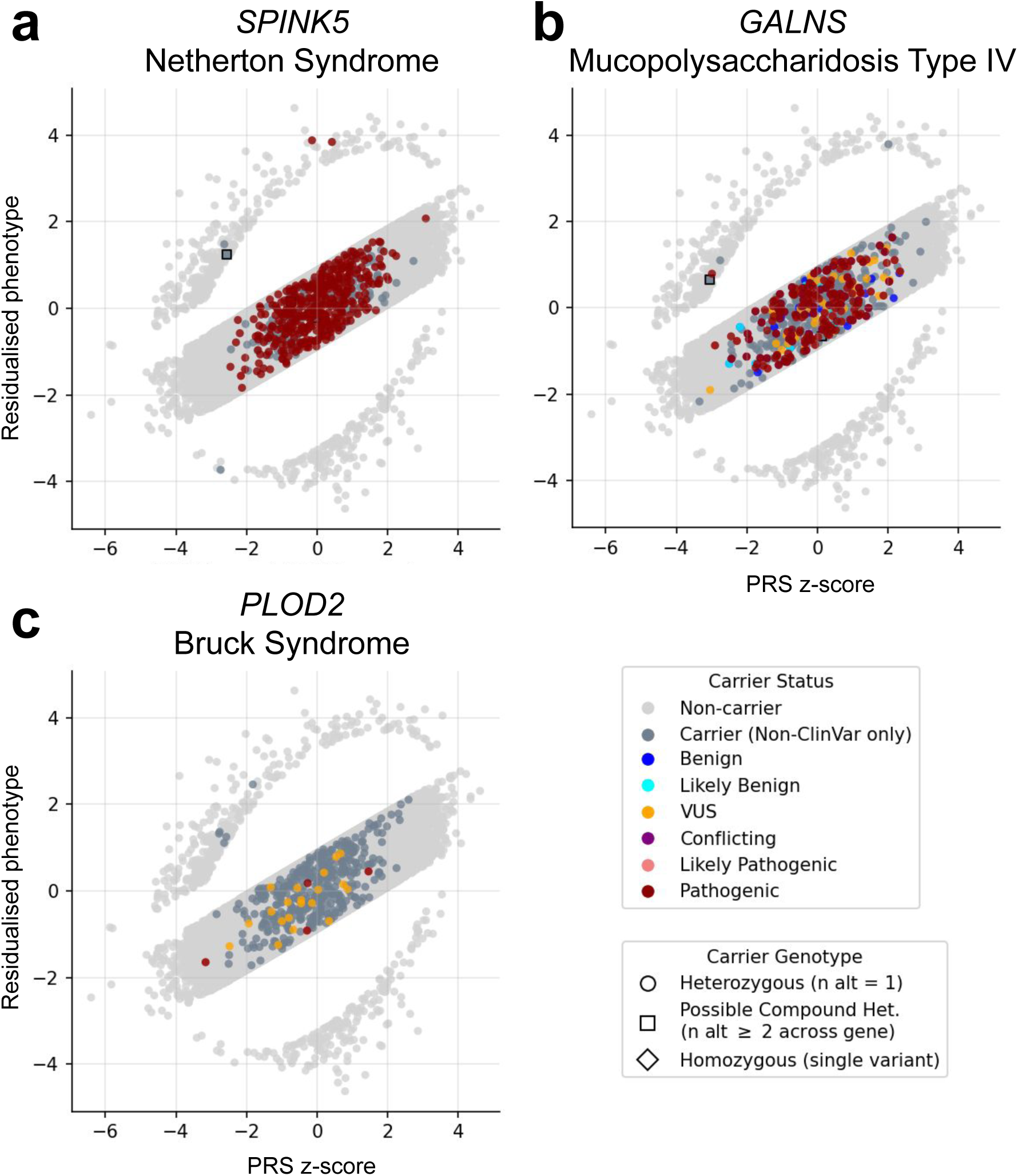
Enrichment of deleterious variants among high outliers for height in genes which cause syndromes associated with short stature. **a**, Enrichment of pLoF variants in *SPINK5*. Enrichment of damaging missense variants in **b**, *GALNS* and **c**, *PLOD2*. Only carriers of the relevant variant consequence category are highlighted (only pLoF or only damaging missense). “Possible Compound Het.” carriers are defined as individuals with two or more alternate alleles of the relevant deleterious variant class (pLoF or damaging missense) across the whole gene, but not at the same variant. Homozygous carriers are those with two alternate alleles at the same variant. ClinVar^81^ annotations for each gene-disorder pairing were obtained from the Open Targets platform^96^.

**Figure S10:**
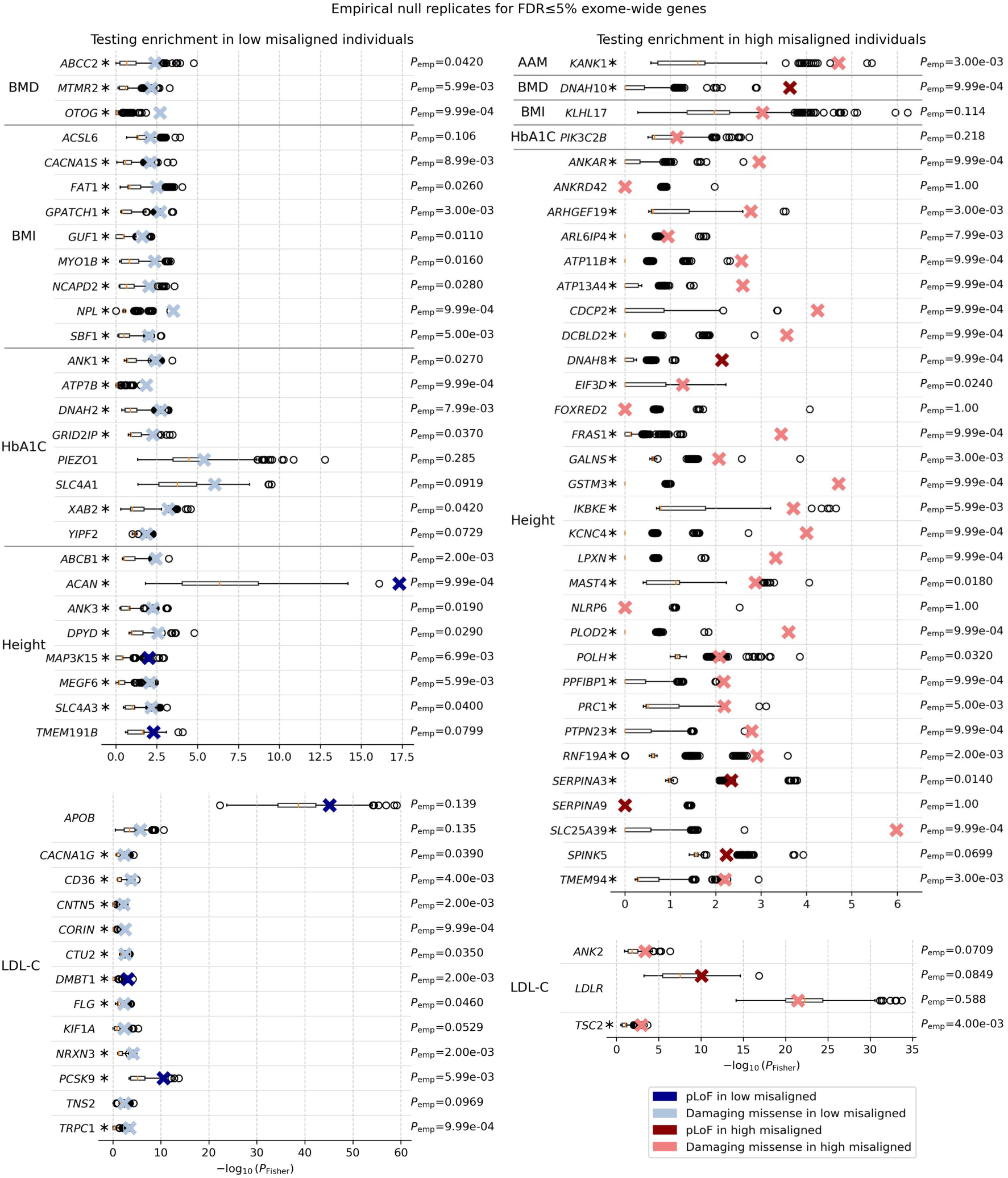
Strength of protective burden association with PRS in disease controls. **a**, Comparison of strength of burden association between original trait (disease case/control status) and protective misaligned effect (effect of pLoF burden on PRS in controls). *P*_misaligned_ is a one-sided *P* -value, with the alternate hypothesis that burden of pLoF variants is associated with higher PRS in disease controls. *P*_original_ is a two-sided *P* -value for the burden association with the original case-control disease trait. Significant genes are defined as having FDR-adjusted one-sided *P*_misaligned_ ≤ 0.05 and are labeled with their gene symbol and, in parentheses, the total gene-level alternate allele count from REGENIE for variants included in the gene burden mask. Canonical monogenic disorder genes are also labeled. Only genes with total alternate allele count ≥ 10 are shown. The total number of significant genes for each disease is shown in parentheses in the title of each plot. **b**, Protective burden effects in FDR-significant genes when not conditioned or conditioned (indicated with “[all]” tag) on all common (MAF *>* 0.1%) QCed imputed variants in or within a 1 Mb radius of the gene tested (Methods). PRS, polygenic risk scores; pLoF, predicted loss-of-function; FDR, false discovery rate

**Figure S9:**
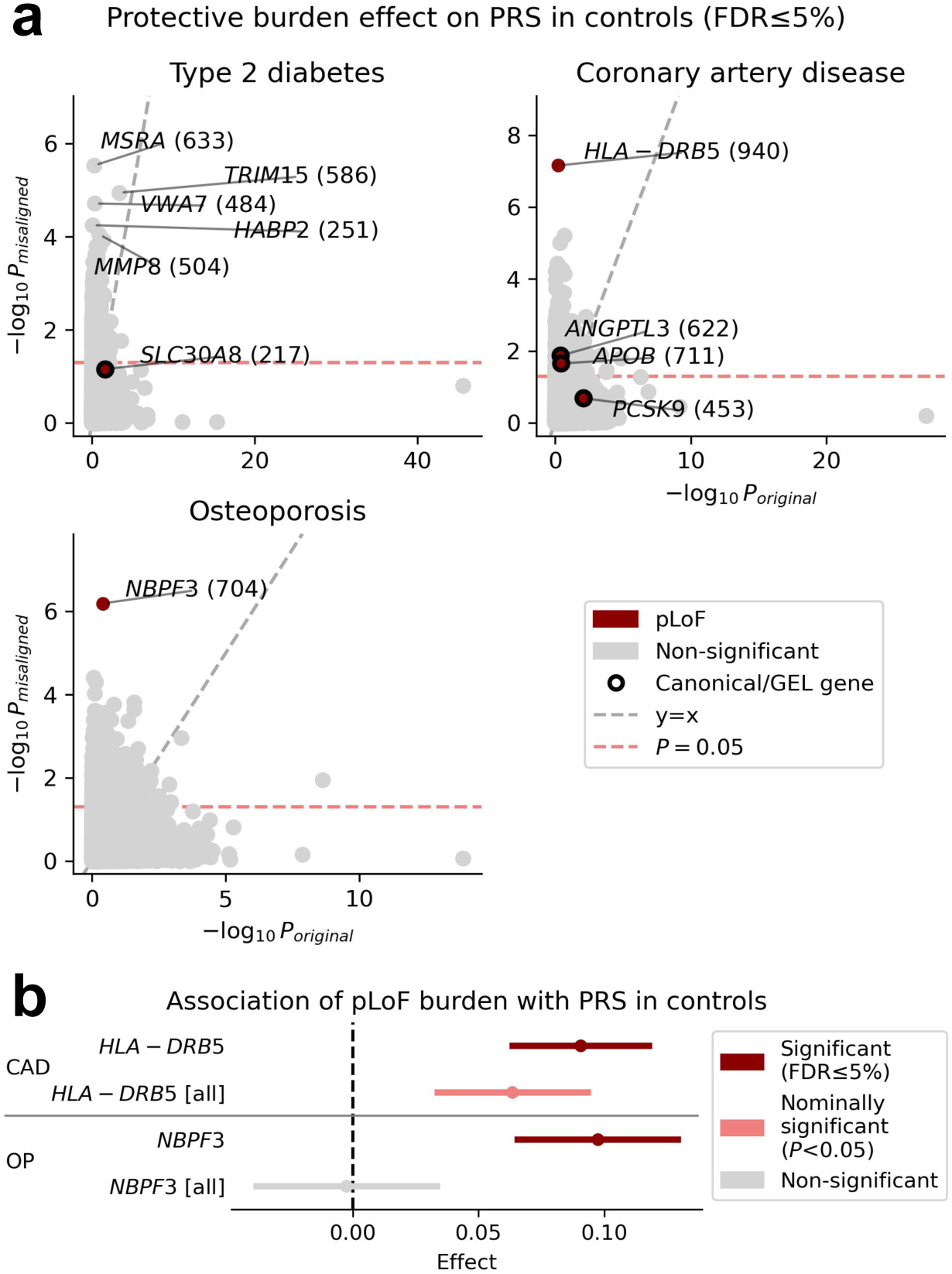
Empirical null replicates for FDR-significant genes nominated by the exome-wide scan. The empirical *P* -value (*P*_emp_, labeled on the right of each plot) of a gene enrichment result indicates how the observed test statistic (one-sided Fisher’s exact test *P* - value) compares against the empirical null distribution of test statistics (Methods). Asterisks immediately to the right of the gene symbol indicate tests where the observed enrichment outperformed the empirical null (*P*_emp_ *<* 0.05). LDL-C is plotted on a different x-axis scale because it tends to have stronger associations.

**Figure S11:**
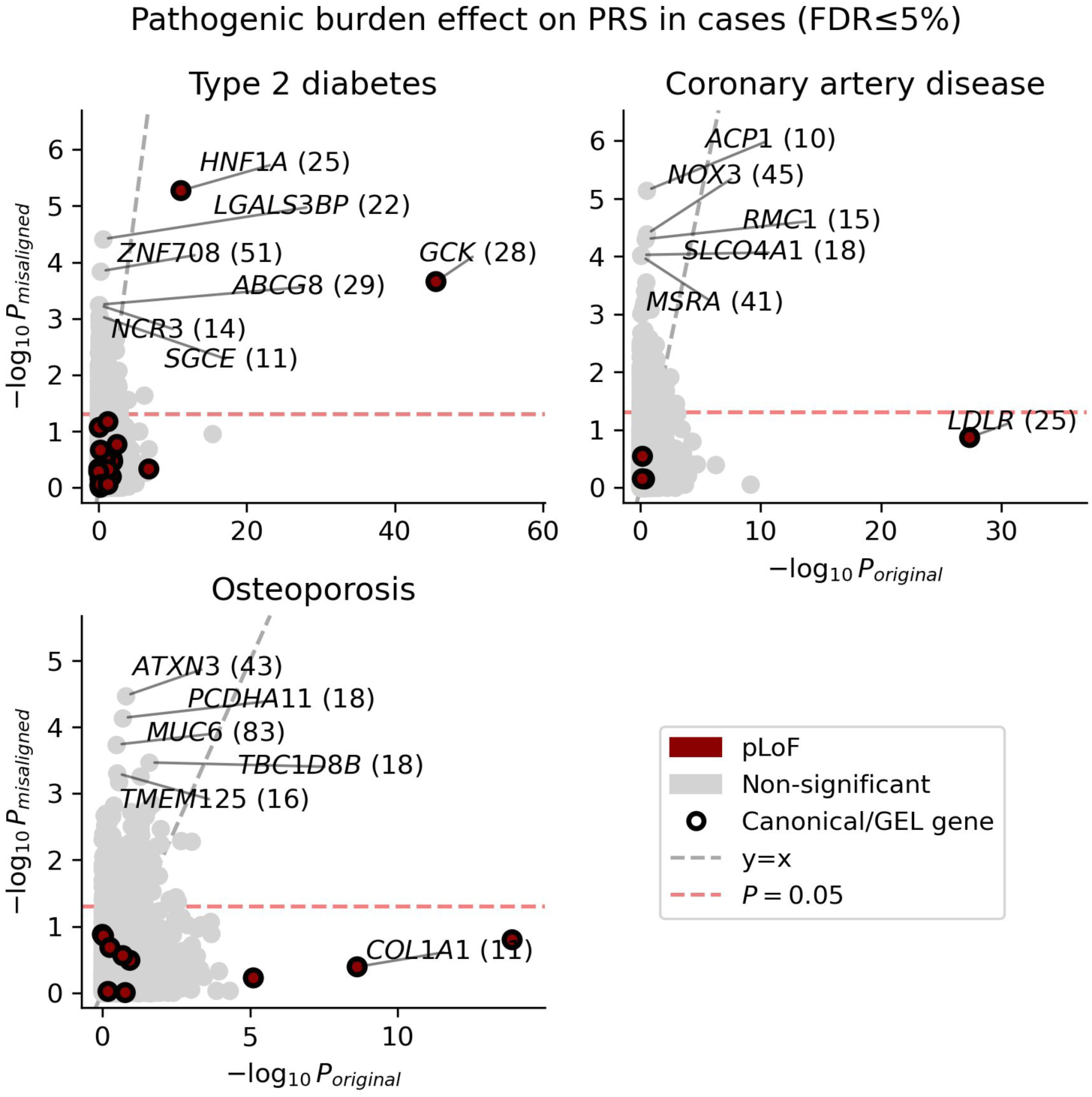
Strength of pathogenic burden association with PRS in disease cases. *P*_misaligned_ is a one-sided *P* -value, with the alternate hypothesis that burden of pLoF variants is associated with lower PRS in disease controls. *P*_original_ is a two-sided *P* -value for the burden association with the original case-control disease trait. Significant genes, of which there were none for the pathogenic burden association testing shown here, are defined as having FDR-adjusted one-sided *P*_misaligned_ ≤ 0.05. The top five genes for each trait with the strongest association strength (highest − log_10_ *P*_misaligned_) are labeled with their gene symbol and, in parentheses, the total gene-level alternate allele count from REGENIE for variants included in the gene burden mask. Canonical monogenic disorder genes are also labeled. Only genes with total alternate allele count ≥ 10 are shown. PRS, polygenic risk scores; pLoF, predicted loss-of-function; FDR, false discovery rate

**Figure S12:**
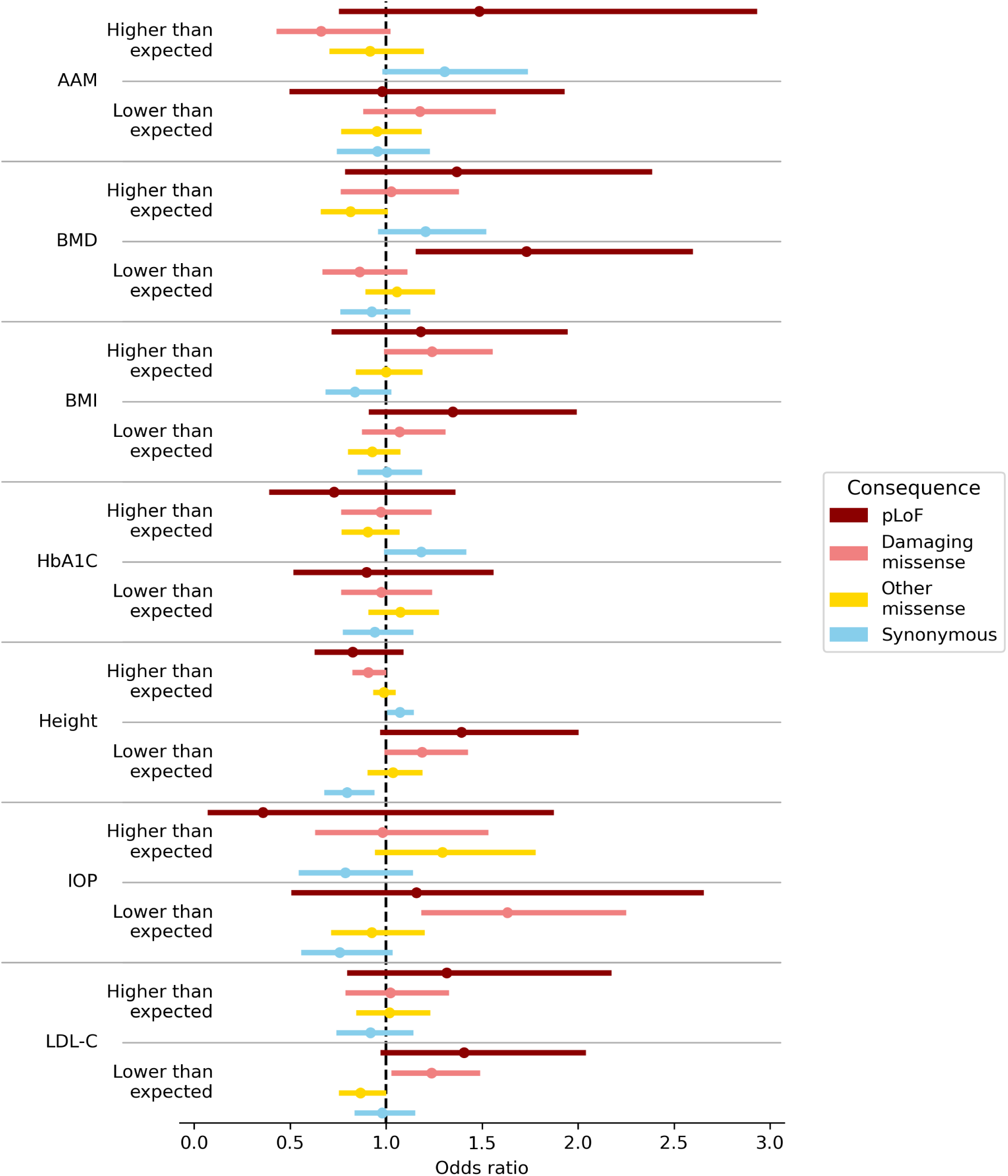
Enrichment of singleton burden in intolerant genes among misaligned individuals for continuous traits. Error bars are two-sided 90% CIs for OR, such that the lower error bar indicates the lower bound of the one-sided 95% CI of the OR from logistic regression of misaligned status against number of singletons, adjusted for total number of singletons (Methods).

**Figure S13:**
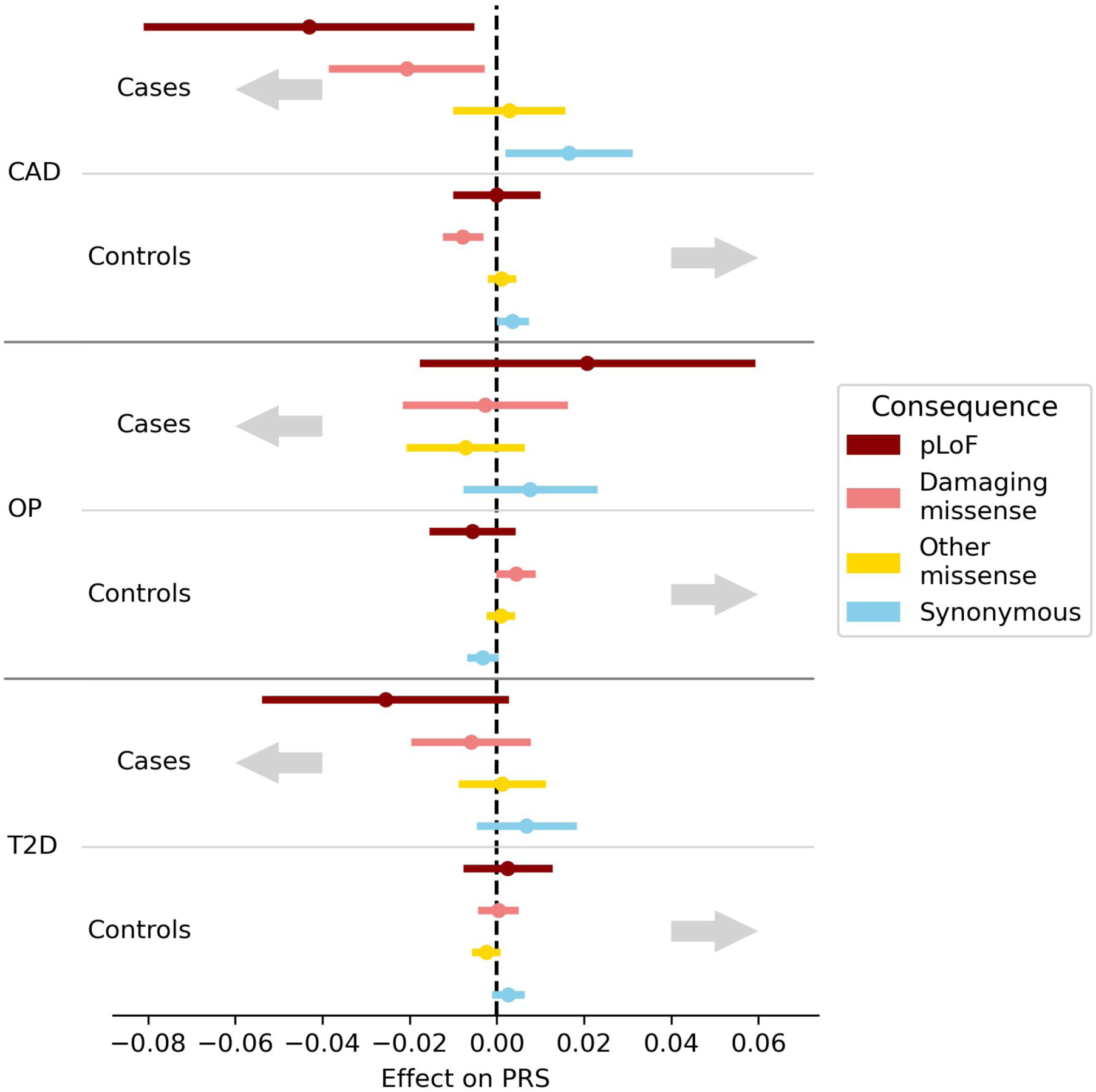
Enrichment of singleton burden in intolerant genes among misaligned individuals for disease traits. Error bars are two-sided 90% CIs for the effect of consequnce-specific singleton burden on PRS (with total number of singletons as a covariate; see Methods), such that the upper bound (when testing cases) and lower bound (when testing controls) correspond to the upper and lower bounds, respectively, of the one-sided 95% CI. Grey arrows indicate the hypothesised direction of effect for the one-sided tests.

## Supplementary tables

For supplementary tables 2-9, see attached supplementary material.

**Table S1:**
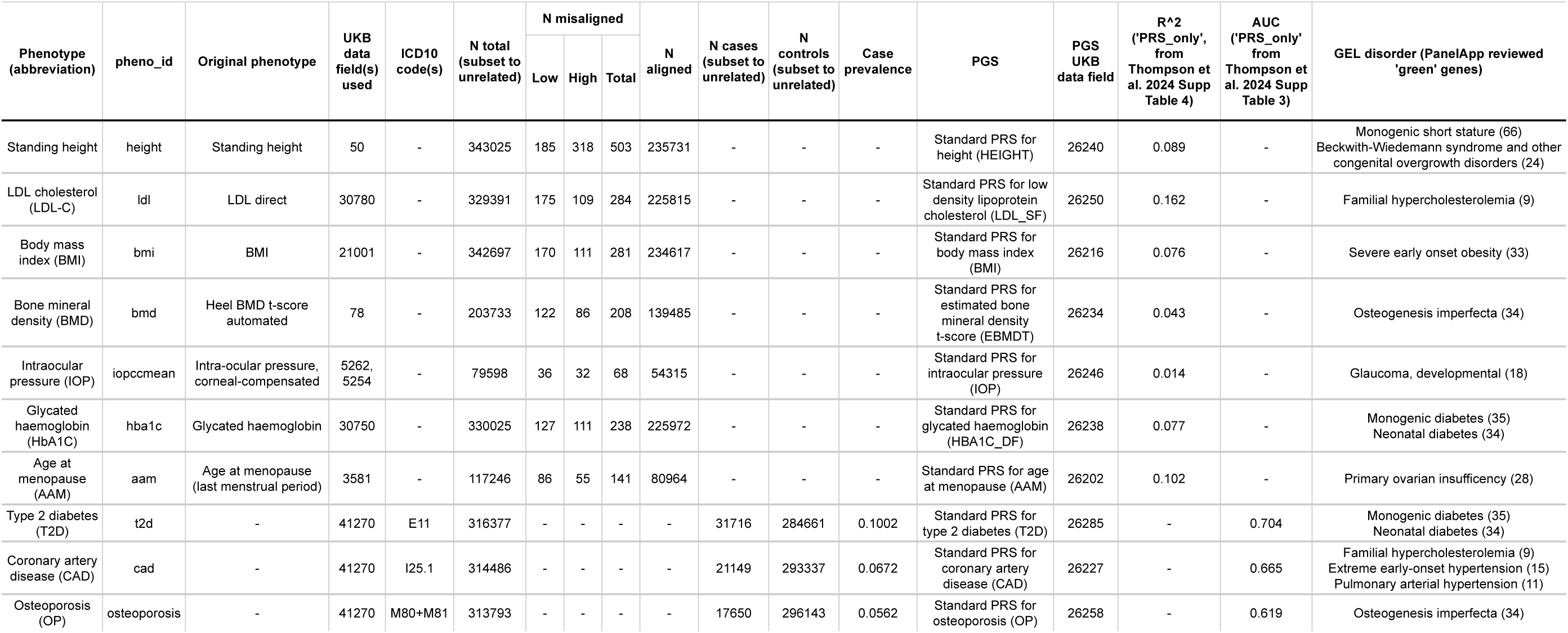
Phenotype/PGS overview.

